# Usual dietary fibre intake in Australia: A comparison of the 2011–12 and 2023 National Nutrition and Physical Activity Surveys

**DOI:** 10.64898/2026.11.14.26368081

**Authors:** Jianan Lu, Rebecca Leech, Matthew Snelson, Francine Z. Marques

## Abstract

**Background & Aims:** Dietary fibre is an essential macronutrient for human health. Whether fibre intake has changed in Australia over the last decade remains unknown. This study estimated usual fibre intake among Australian children and adults in 2023 and compared it with 2011–12.

**Methods:** We used data from the 2011–12 (n=12,153) and 2023 (n=11,198) National Nutrition and Physical Activity Surveys with repeat 24-hour recalls. Usual intake was estimated with the National Cancer Institute method and compared with the Adequate Intake (AI) and the Suggested Dietary Target (SDT) levels. Fibre density (g/1,000 kcal) was modelled jointly with energy.

**Results:** Mean usual fibre intake in adults decreased by 2.1 g/day (95% confidence interval 1.6-2.6), from 22.9 to 20.8 g/day. The proportion meeting the AI fell from 24.6% to 17.2%, and SDT from 11.4% to 7.5%. Declines occurred in both sexes and were largest among adults aged 51 years and over. Fibre density fell by 3.2% to 10.7 g/1,000 kcal. Intake in children was stable (19.7 and 20.0 g/day), but only 21.8% of adolescents aged 14–18 years met the AI. In 2023, intake was significantly lower in adults from the most disadvantaged areas and in those with hypertension or obesity.

**Conclusions:** Usual fibre intake among Australian adults declined over the last decade, reflecting both lower energy intake and less fibre-rich food choices. Strategies to improve the availability, affordability and uptake of fibre-rich foods are needed, particularly for adolescents, older adults and socioeconomically disadvantaged groups.

## 1. Introduction

Dietary fibre is a key component of a healthy diet. Higher intakes are consistently associated with a lower risk of non-communicable chronic disease [1]. Comparing the highest with the lowest intakes, prospective cohort studies show a lower incidence of coronary heart disease, stroke, type 2 diabetes, and colorectal cancer, and 15–30% lower all-cause and cardiovascular mortality [1]. The greatest risk reduction is observed at intakes of 25–29 g/day, with dose-response data suggesting further benefits at higher intakes [1]. Meanwhile, diets low in whole grains, a major source of fibre, accounted for an estimated 3 million deaths worldwide in 2017 – similar to high sodium intake – and ranked among the top three leading dietary risks for death and disability [2]. Therefore, monitoring whether populations achieve recommended fibre intake is central to the prevention of non-communicable disease.

Despite its established health benefits, dietary fibre intake remains below dietary targets across most populations [2]. National dietary surveys in Europe and North America consistently report low intakes, with fewer than a quarter of adults, and as few as 4% in some countries, meeting country-specific recommendations [3–6]. In Australia, the 2011–12 National Nutrition and Physical Activity Survey (NNPAS) estimated that only 28% of adults met the Adequate Intake (AI, levels that support normal laxation) and fewer than 20% met the Suggested Dietary Target (SDT, levels to reduce the long-term risk of chronic disease) [7]. Attainment also varied across the population, with adolescents aged 14–18 years, young adults aged 19–30 years, men and people of lower socioeconomic status least likely to meet fibre targets [7].

Yet whether population fibre intake has changed over time remains unclear. National dietary surveys are often conducted infrequently, while advances in dietary assessment methods can complicate comparisons across survey years [8–10]. Furthermore, estimates based on a single 24-hour recall may misrepresent usual intake distributions and the proportion of individuals meeting dietary targets [11,12]. These limitations leave important gaps in understanding long-term trends in fibre intake and whether changes differ across population groups, particularly those at elevated cardiovascular risk.

Australia’s 2023 NNPAS, the first national nutrition survey in more than a decade, provides a timely opportunity to address these evidence gaps [13]. Using nationally representative data from the 2011–12 and 2023 surveys, we aimed to (1) estimate usual dietary fibre intake among children and adults in 2023 and the proportions meeting AI and SDT; (2) assess changes in usual intake and target attainment over time; and (3) examine differences in intake across demographic, socioeconomic and cardiovascular risk groups.

## 2. Methods

### 2.1 Survey design and participants

The 2023 NNPAS was a nationally representative, cross-sectional household survey conducted by the Australian Bureau of Statistics (ABS) between January 2023 and March 2024 as part of the Intergenerational Health and Mental Health Study [13]. The survey included usual residents aged 2 years and over living in private dwellings in urban and rural areas of all Australian states and territories, excluding very remote areas and discrete Aboriginal and Torres Strait Islander communities [13]. Households were randomly selected, and one adult aged 18 years or over and, where applicable, one child aged 2–17 years were randomly selected from each participating household. A total of 8,817 households responded, representing a household response rate of 54.9%, and dietary data were collected from 11,199 participants [13].

For children aged 2–14 years, a parent or guardian provided the survey responses; adolescents aged 15–17 years could respond themselves with parental or guardian consent, or a parent or guardian responded on their behalf. Survey weights supplied by the ABS accounted for participants’ probabilities of selection and were calibrated to the Australian in-scope population in June 2023 and the distribution of dietary recall days [13].

The 2011–12 NNPAS, conducted as part of the 2011–13 Australian Health Survey, used a similar design and scope, described previously [7,14]. It was conducted between May 2011 and June 2012; 9,519 households responded (77.0%), yielding 12,153 participants aged 2 years and over [14].

The ABS conducted both surveys under the Census and Statistics Act 1905; the ABS obtained ethics approval only for the separate biomedical component of the 2011–12 survey [13,14]. This secondary analysis used de-identified ABS Basic Microdata, accessed by the authors as approved users. The Monash University Human Research Ethics Committee granted ethics approval (project ID 53260).

### 2.2 Sociodemographic and health characteristics

All characteristics were taken and harmonised from the ABS microdata files across the two surveys (Table S1) [13,14]. Records with missing, not stated or not applicable values were excluded from estimates for that variable only. Education, labour force status, marital status, living alone, smoking, hypertension and diabetes or high sugar levels were reported for adults aged 19 years and over. Variable names and category labels follow the ABS data item lists for each survey.

Age was grouped to match the age bands of the fibre reference values: 2–3, 4–8, 9–13 and 14–18 years for children, and 19–30, 31–50, 51–70 and 71 years and over for adults [7]. Sex was recorded as sex at birth in 2023 and as sex in 2011–12. Country of birth was recorded in three ABS categories (Australia, Main English-Speaking Countries and Other), and the share born in Australia is reported. Area-level socioeconomic status was measured with the Index of Relative Socio-economic Disadvantage (IRSD), one of the ABS Socio-Economic Indexes for Areas (SEIFA). National IRSD quintiles were assigned at Statistical Area Level 1, using the 2011 index for the 2011–12 survey and the 2021 index for the 2023 survey [15,16]. Individual-level socioeconomic status was measured with equivalised household income, which adjusts total household income for household size and composition; ABS deciles were combined in pairs to form quintiles. Remoteness was classified using the ABS Remoteness Areas of the Australian Statistical Geography Standard (2011 and 2021 editions) into three categories in the ABS Basic Microdata: Major Cities of Australia, Inner Regional Australia, and Other (outer regional, remote, and very remote areas combined).

Highest non-school qualification was grouped as bachelor degree or above (Postgraduate Degree, Graduate Diploma and Graduate Certificate, and Bachelor Degree), certificate or diploma (Advanced Diploma and Diploma, Certificate III and IV, Certificate I and II, and Certificate n.f.d.), or no non-school qualification; level not determined was excluded. Labour force status was classified as employed, unemployed or not in the labour force. Marital status was the ABS social marital status (married or not married), which the ABS derived differently in the two surveys. Living alone was defined as a household of one person.

Current smokers comprised current daily, weekly, and less-than-weekly smokers in 2011–12, and current daily smokers and other current smokers in 2023. Ex-smokers and people who had never smoked (2011–12) and current non-smokers (2023) were classified as not current smokers, because the 2023 survey did not separate them. Trained interviewers measured height and weight, and body mass index (BMI) was calculated as weight (kg) divided by height squared (m^2^). Adults were classified using World Health Organisation cut-offs as underweight (<18.5 kg/m2), normal weight (18.5–24.9 kg/m2), overweight (25.0–29.9 kg/m2), or obese (≥30.0 kg/m^2^) [17]. Children were classified with the International Obesity Task Force age- and sex-specific cut-offs [18,19]. Underweight combined the ABS thinness grades 1–3, and obese combined obesity classes 1–3. In 2023, the ABS imputed BMI for participants who were not measured; in 2011–12, unmeasured participants were treated as missing. Pregnant women were not assigned a BMI category in either survey.

In 2023, hypertension was defined with the ABS-derived variable for self-reported hypertension or high measured blood pressure, defined by the ABS as a systolic pressure of 140 mmHg or above and/or a diastolic pressure of 90 mmHg or above [20]. The ABS imputed blood pressure by hot-deck imputation for participants who were not measured [21]. In 2011–12, hypertension was defined as a measured systolic blood pressure of 140 mmHg or above or diastolic blood pressure of 90 mmHg or above, or a self-reported current diagnosis of hypertensive disease. Pregnant women were excluded from the hypertension estimates in both surveys.

Self-reported diabetes or hyperglycaemia was defined from the ABS item for age first told had diabetes or high sugar levels, which has the same categories in both surveys. Participants with an age band (under 25 to 65 and over) or a response of not known were classified as having been told, and those coded not applicable as not told. The item includes people told they have high sugar levels without a diagnosis of diabetes, so it does not identify current diabetes.

### 2.3 Dietary assessment

In 2023, dietary intake was assessed using Intake24, a multiple-pass 24-hour dietary recall system developed and validated in the United Kingdom [22]. The ABS adapted Intake24 for use in Australia in collaboration with Monash University and Food Standards Australia New Zealand (FSANZ) [13]. The first recall was interviewer-guided and completed during the face-to-face survey interview. Participants were subsequently invited to complete a second recall at least 8 days later, either online or during a second face-to-face interview; 9,032 participants completed the second recall [13]. Reported foods and beverages were linked to the AUSNUT 2023 food and nutrient database developed by FSANZ.

In 2011–12, trained interviewers collected the first 24-hour recall in a face-to-face interview using the Automated Multiple-Pass Method (AMPM), adapted from the United States Department of Agriculture instrument. A second recall was completed by telephone at least 8 days later by 7,735 participants (64%) [14]. Based on the recorded recall dates, the median interval between the two recalls was 17 days (interquartile range 13–24). Reported foods were linked to the AUSNUT 2011–13 database developed by FSANZ.

### 2.4 Dietary fibre intake and reference values

Dietary fibre intake (g/day) on each recall day was derived by the ABS from all foods and beverages reported, using AUSNUT 2011–13 for the 2011–12 survey and AUSNUT 2023 for the 2023 survey [13,14]. Fibre from dietary supplements was excluded because supplement fibre was recorded only for the first recall in 2023 and the AI was derived from fibre intakes from food [13,23]. Dietary fibre is defined in the Australia New Zealand Food Standards Code as the fraction of the edible parts of plants, their extracts or synthetic analogues that resists digestion and absorption in the small intestine. It includes polysaccharides, oligosaccharides with a degree of polymerisation greater than 2, and lignins [23].

Usual fibre intakes were compared with the Nutrient Reference Values for Australia and New Zealand [23]. The AI is 14 g/day for children aged 2–3 years and 18 g/day for those aged 4–8 years. For ages 9–13 years, it is 24 g/day for boys and 20 g/day for girls; for ages 14–18 years, it is 28 g/day for boys and 22 g/day for girls. For adults aged 19 years and over, the AI is 30 g/day for men and 25 g/day for women. The AI is based on median intakes in Australia and New Zealand, with an allowance for resistant starch, rather than on a physiological requirement [23]. All women were assessed against the non-pregnant AI of 25 g/day.

For adults, intakes were also compared with the SDT for chronic disease prevention: 38 g/day for men and 28 g/day for women. The SDT corresponds to the 90^th^ centile of population intake and was set to reduce the risk of coronary heart disease [23]. No SDT is defined for children. The proportion meeting each reference value was estimated as the proportion of the usual intake distribution at or above that value, within each age and sex group. Because the AI is not a requirement, usual intakes below the AI cannot be interpreted as inadequate [24].

Fibre density was expressed as grams per 1,000 kcal of energy intake, with energy including the contribution from fibre (1 kcal=4.184 kJ). Fibre density was compared with 14 g/1,000 kcal, the energy-based benchmark used to set the United States AI for fibre [25].

### 2.5 Usual intake estimation

Usual fibre intake distributions were estimated with the National Cancer Institute (NCI) method. The method uses repeat 24-hour recalls in a subsample to separate within-person (day-to-day) variation from between-person variation [11,12]. Fibre was treated as a nutrient consumed daily, so a one-part model of the amount consumed was used. Models were fitted in R with the ncimultivar package (version 1.1.2), which implements the NCI method using Markov chain Monte Carlo (MCMC) estimation [26,27].

Daily fibre intakes were Box-Cox transformed, with the transformation parameter estimated from Day 1 recalls. Transformed intakes were modelled with a linear mixed-effects model containing a person-level random effect (between-person variation) and a residual term (within-person variation). Separate models were fitted for each survey, because day-to-day variability and the Box-Cox transformation could differ between surveys. Separate models were also fitted for adults (19 years and over) and children (2–18 years). For participant *i* on recall day *j*, the reported fibre intake (*R_ij_*) was transformed and modelled as:

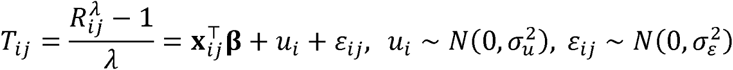

where *λ* is the Box-Cox parameter, *X_ij_* is the vector of covariates with coefficients **β**, *u_i_* is the person-level random effect with between-person variance 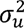, and *ε_ij_* is the within-person error with within-person variance 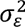. The two terms are independent. Usual intake is the long-run average daily intake of a person on the original scale:

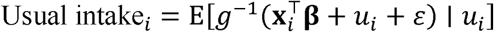

where *g*^-1^ is the inverse Box-Cox transformation. Because the back-transformation is non-linear, we evaluated this expectation by simulation, as described below.

The adult model included sex, area-level socioeconomic quintile and age, standardised to a mean of 0 and a variance of 1. The children’s model included a term for each of the eight sex-by-age groups that define the AI (2–3, 4–8, 9–13 and 14–18 years), and socioeconomic quintile. In children, between- and within-person variances were shared across age groups, because only about half of 2–3-year-olds (147 of 301) completed two recalls in 2023. All models included indicators for the second recall and for weekend consumption (Saturday or Sunday, based on the day the food was eaten). Each model was run for 3,000 MCMC iterations, with the first 1,000 discarded as burn-in and every second iteration retained. Convergence of each main model was assessed with the Gelman–Rubin statistic across five chains, with values below 1.1 taken to indicate convergence [28].

Usual intakes were simulated for every participant from the fitted model, using 200 random-effect draws per participant. The second-recall indicator was set to zero, and simulated weekday and weekend intakes were averaged with weights of 5 and 2. Simulated intakes were back-transformed to grams per day, with a lower bound of half the smallest non-zero Day 1 intake. Means, percentiles and the proportions at or above the AI and SDT were then calculated from the weighted simulated distribution, overall and within each subgroup. Extreme intakes were identified with the package’s default rule: values more than three interquartile ranges below the 25^th^ or above the 75^th^ percentile on the transformed scale.

These values were reviewed but not Winsorised. In the adult models, 32 (2011–12) and 42 (2023) Day 1 values met this rule; in the children’s models, 5 and 9 did. Three implausible daily fibre values above 120 g/day in 2023 (one Day 1 and two Day 2) were removed before modelling.

For fibre density, fibre and energy (kcal; kilojoules divided by 4.184) were modelled jointly in a bivariate model of two daily-consumed nutrients, in which each followed the model above and their person-level random effects were allowed to be correlated. The Box-Cox parameter was estimated separately for each variable, and the covariates, weighting, MCMC settings, convergence checks, and simulation matched those in the fibre-only models. Participants with no valid Day 1 energy value were excluded (adults: 4 in 2011–12 and 8 in 2023; children: none in 2011–12 and 1 in 2023). Usual fibre density was calculated for each simulated participant as 1,000 × usual fibre intake ÷ usual energy intake (g/1,000 kcal), applying the ratio to usual intakes rather than daily ratios, as recommended [26,27]. The population estimate of fibre density was the ratio of weighted mean usual fibre to weighted mean usual energy; the proportion at or above 14 g/1,000 kcal was also calculated.

Sampling variability was estimated by refitting each model 60 times, once with each of the 60 ABS replicate weights, and re-simulating usual intakes for every refit with replicate-specific random seeds. The standard error (SE) of each estimate 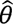 was 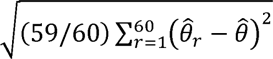, where 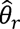 is the estimate from replicate *r*, and 95% confidence intervals (CIs) were the estimate ± 1.96 SE. The SE of a change between surveys was 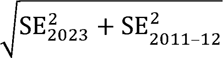, because the two samples are independent.

### 2.6 Statistical analysis

The study was reported according to the Strengthening the Reporting of Observational Studies in Epidemiology–nutritional epidemiology (STROBE-nut) checklist (Table S2) [29]. All estimates used the Day 1 person weight supplied by the ABS, including in the usual-intake models, where it was applied to both recalls. The Day 2 weight, which adjusts for non-response among participants who completed a second recall, was not used. Applying it would restrict the analysis to participants with two recalls and exclude those with Day 1 data only (36% in 2011–12 and 19% in 2023). The proportion completing a second recall differed between surveys (64% and 81%). Weights were not recalibrated after removing the three implausible values.

The outcomes were mean usual fibre intake; the 5^th^, 25^th^, 50^th^, 75^th^, and 95^th^ percentiles; the proportion at or above the AI (adults and children) and the SDT (adults); and fibre density. These were estimated overall and by sex, age group, sex-by-age group and socioeconomic quintile, with 95% CIs from the replicate-weighted jackknife. Differences between surveys were calculated as 2023 minus 2011–12, and a difference was considered statistically significant when its 95% CI excluded zero. The analysis was descriptive, and no adjustment was made for multiple comparisons. Observed Day 1 intakes were analysed alongside usual intakes, with the same weights, groups, and jackknife, to show how much of the spread in intakes, and in the proportion meeting the AI and SDT, reflects day-to-day variation (Table S3).

Subgroup analyses were restricted to adults and covered socioeconomic and cardiovascular risk groups. Estimates by area-level socioeconomic quintile came from the main usual-intake model, which included quintile as a covariate. BMI category (underweight, normal weight, overweight and obese), hypertension, and self-reported diabetes or hyperglycaemia were not covariates in the main model. For these, the usual-intake model was refitted separately within each group and survey, with the same covariates, MCMC settings, simulation and jackknife as the main model. Subgroup results are descriptive, and differences between groups were not formally tested. Analyses were done in R version 4.3.3 with ncimultivar version 1.1.2, with fixed random-number seeds for all simulations and refits.

## 3. Results

### 3.1 Participants and comparability of the surveys

A total of 12,153 participants in 2011–12 and 11,199 in 2023 completed a Day 1 recall (Figure S1). In 2023, one participant with an implausible Day 1 fibre intake was excluded and two implausible Day 2 values were set to missing; all three values were above 120 g/day. No participants were excluded in 2011–12. The analytic sample therefore comprised 12,153 participants in 2011–12 and 11,198 in 2023. Completion of the second recall was higher in 2023 (n=9,029; 80.6%) than in 2011–12 (n=7,735; 63.6%).

The two samples had similar distributions of sex, area-level socioeconomic disadvantage, equivalised household income, remoteness, and labour force status (Table 1). The 2023 sample was slightly older, with 10.9% aged 71 years and over compared with 8.4% in 2011–12. Fewer participants were born in Australia in 2023 (67.9% in 2023 versus 73.7% in 2011-12). Among adults, the proportion with a bachelor’s degree or above rose from 25.7% to 37.0%, and current smoking fell from 17.7% to 11.1%. The prevalence of obesity (28.6% versus 22.0%), self-reported diabetes or hyperglycaemia (13.7% versus 10.2%) and hypertension (32.0% versus 30.8%) was higher in 2023.

**Table 1.** Characteristics of participants aged 2 years and over, National Nutrition and Physical Activity Survey (NNPAS) 2011–12 and 2023.

| **Characteristic** | **2011–12 (n = 12,153)<br>n (%)** | **2023 (n = 11,198)<br>n (%)** |
|---|---|---|
| **Sociodemographic characteristics** |  |  |
| **Age group (years)** |  |  |
| 2–3 | 464 (2.6) | 301 (2.3) |
| 4–8 | 789 (6.4) | 738 (6.2) |
| 9–13 | 787 (7.1) | 688 (6.4) |
| 14–18 | 772 (6.1) | 725 (6.4) |
| 19–30 | 1,592 (18.0) | 1,198 (16.6) |
| 31–50 | 3,565 (29.1) | 3,013 (28.1) |
| 51–70 | 2,906 (22.3) | 2,793 (23.1) |
| 71 and over | 1,278 (8.4) | 1,742 (10.9) |
| **Sex** |  |  |
| Male | 5,702 (49.7) | 5,314 (49.5) |
| Female | 6,451 (50.3) | 5,884 (50.5) |
| **Born in Australia** | 9,157 (73.7) | 7,902 (67.9) |
| **Area-level socioeconomic disadvantage (IRSD quintile)** |  |  |
| Quintile 1 (most disadvantaged) | 2,238 (17.9) | 1,984 (18.1) |
| Quintile 2 | 2,464 (19.8) | 2,191 (20.4) |
| Quintile 3 | 2,432 (20.7) | 2,335 (21.1) |
| Quintile 4 | 2,177 (18.9) | 2,165 (20.8) |
| Quintile 5 (least disadvantaged) | 2,842 (22.7) | 2,523 (19.6) |
| **Equivalised household income quintile<sup>a</sup>** |  |  |
| Quintile 1 (lowest) | 2,179 (18.9) | 1,858 (17.4) |
| Quintile 2 | 2,028 (18.5) | 1,812 (17.6) |
| Quintile 3 | 2,134 (21.5) | 2,193 (21.5) |
| Quintile 4 | 2,400 (21.6) | 2,219 (22.7) |
| Quintile 5 (highest) | 2,159 (19.6) | 2,109 (20.7) |
| **Remoteness** |  |  |
| Major cities | 7,788 (70.7) | 7,158 (72.5) |
| Inner regional | 2,376 (19.7) | 2,186 (19.2) |
| Outer regional and remote | 1,989 (9.6) | 1,854 (8.3) |
| **Highest non-school qualification<sup>b,c</sup>** |  |  |
| Bachelor's degree or above | 2,385 (25.7) | 3,129 (37.0) |
| Certificate or diploma | 3,177 (35.5) | 3,057 (34.7) |
| None | 3,636 (38.8) | 2,407 (28.3) |
| **Labour force status<sup>b</sup>** |  |  |
| Employed | 5,949 (66.9) | 5,405 (66.2) |
| Unemployed | 208 (2.5) | 122 (2.0) |
| Not in the labour force | 3,184 (30.6) | 3,219 (31.8) |
| **Married<sup>b</sup>** | 4,958 (59.6) | 4,829 (63.4) |
| **Living alone<sup>b</sup>** | 2,535 (14.0) | 2,595 (16.0) |
| **Health characteristics** |  |  |
| **Current smoker<sup>b</sup>** | 1,769 (17.7) | 1,006 (11.1) |
| **Body mass index category<sup>d</sup>** |  |  |
| Underweight | 239 (2.6) | 281 (2.4) |
| Normal weight | 4,273 (42.8) | 4,084 (38.1) |
| Overweight | 3,310 (32.7) | 3,445 (30.8) |
| Obese | 2,356 (22.0) | 3,235 (28.6) |
| **Self-reported hypertension<sup>b,e</sup>** | 2,765 (30.8) | 3,169 (32.0) |
| **Self-reported diabetes or hyperglycaemia<sup>b</sup>** | 1,085 (10.2) | 1,268 (13.7) |

Values are unweighted n (weighted %); percentages are weighted to the Australian population and exclude missing values. Definitions and differences in measurement between the surveys are described in Section 2.2 and Table S1. IRSD, Index of Relative Socio-economic Disadvantage.
<sup>a</sup> Excludes participants with household income not stated or not known (2011–12, 1,253; 2023, 1,007).
<sup>b</sup> Adults aged 19 years and over (2011–12, n = 9,341; 2023, n = 8,746).
<sup>c</sup> Excludes participants whose qualification level could not be determined (2011–12, 143; 2023, 153).
<sup>d</sup> Excludes pregnant women and participants without a usable measurement (2011–12, 1,975; 2023, 153).
<sup>e</sup> Excludes pregnant women and, in 2011–12, adults with neither a blood pressure reading nor a self-reported diagnosis (2011–12, 1,215; 2023, 74).

### 3.2 Day-to-day variation in fibre intake

Like most of the diet, fibre intake varied from day to day within the same person. In adults, the within-person variance was 1.44 times the between-person variance in 2011–12 and 1.39 times in 2023. In children, the ratios were 2.28 and 2.12. A single recall therefore combines a person’s habitual intake with the particular day sampled; the NCI method separates the two.

Among adults with two recalls, the correlation between Day 1 and Day 2 fibre intake was 0.43 in 2011–12 and 0.45 in 2023. In both surveys, only 38% of adults fell in the same intake quartile on both days. Of those in the top quartile on Day 1, 48% were also in the top quartile on Day 2, and about 5% moved between the bottom and top quartiles (Figure S2). A single day therefore placed many people above or below their habitual intake, in both directions.

Removing day-to-day variation did not change mean intake, and it did not account for the low attainment. In 2023, the mean adult intake was 20.7 g/day on Day 1 and 20.8 g/day as usual intake. Usual intake was less dispersed than single-day intake, so fewer adults, not more, were estimated to meet the reference values: 17.2% met the AI and 7.5% the SDT based on usual intake, compared with 21.7% and 13.1% based on Day 1 alone (Table S3). In children, the 5^th^ to 95^th^ percentile range narrowed from 7.2–38.2 g/day on Day 1 to 11.4–30.9 g/day as usual intake. Usual intake was used for all remaining analyses.

### 3.3 Usual fibre intake and target attainment in 2023

In 2023, the mean usual fibre intake of adults was 20.8 g/day (95% CI = 20.5 to 21.1), 22.2 g/day in men and 19.5 g/day in women (Table 2). Most adults fell below the reference values (Figure 1). Only 17.2% (95% CI = 15.7 to 18.7) met the AI and 7.5% (95% CI = 6.4 to 8.6) met the SDT. Attainment was lowest for the SDT in men (3.7%). Mean intake and AI attainment varied little across adult age groups (20.5 to 21.0 g/day; 16.5% to 17.5%).

**Figure 1.**
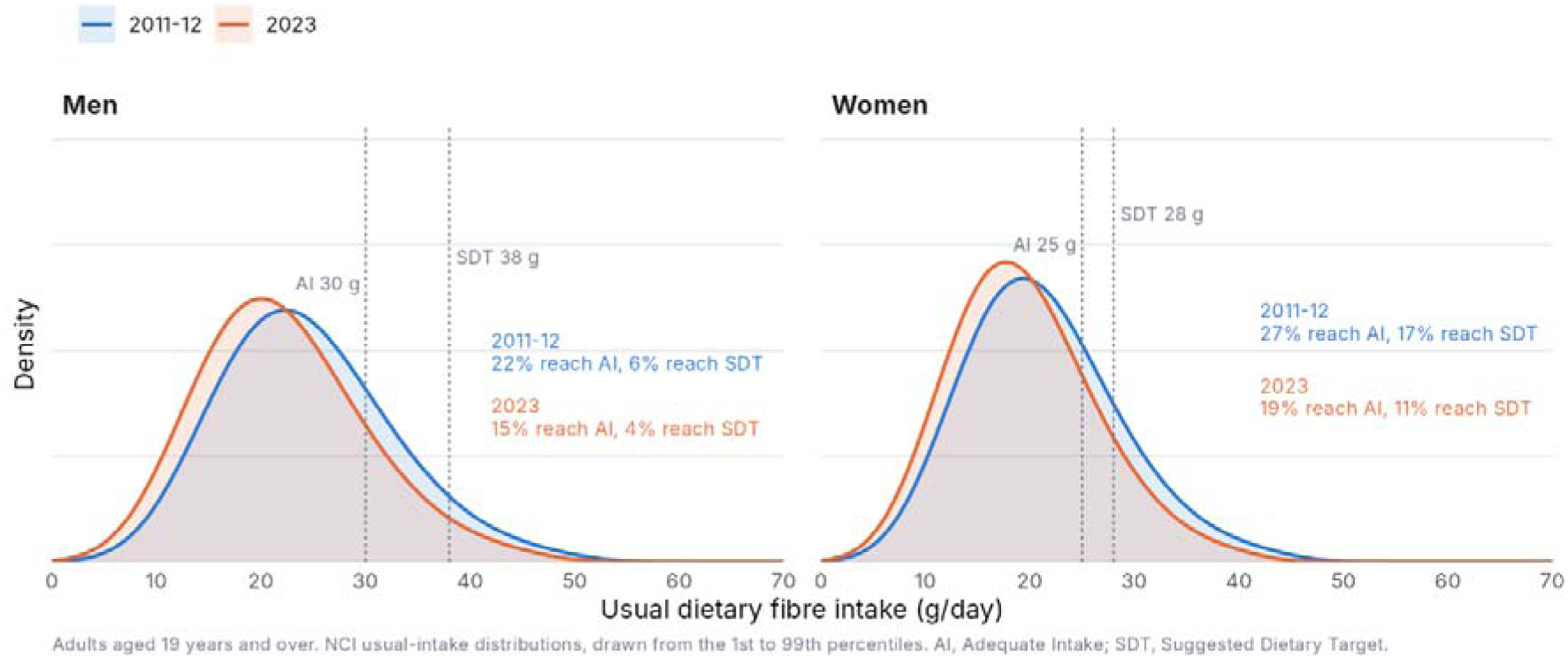
Distribution of usual dietary fibre intake in Australian men and women aged 19 years and over, NNPAS 2011–12 and 2023. ^a^ Dashed lines mark the Adequate Intake (AI; 30 g/day for men and 25 g/day for women) and the Suggested Dietary Target (SDT; 38 g/day for men and 28 g/day for women).

**Table 2.**
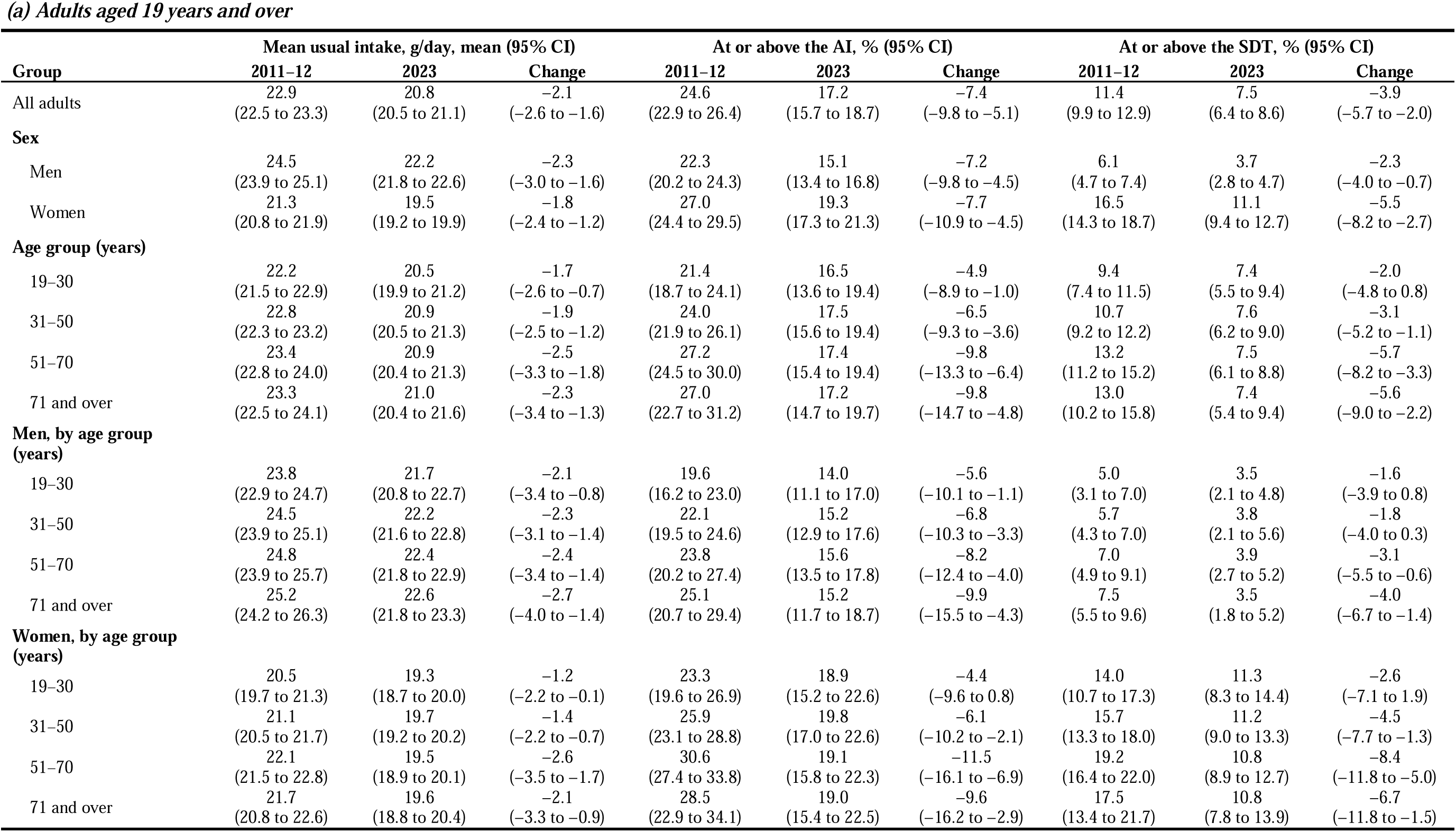
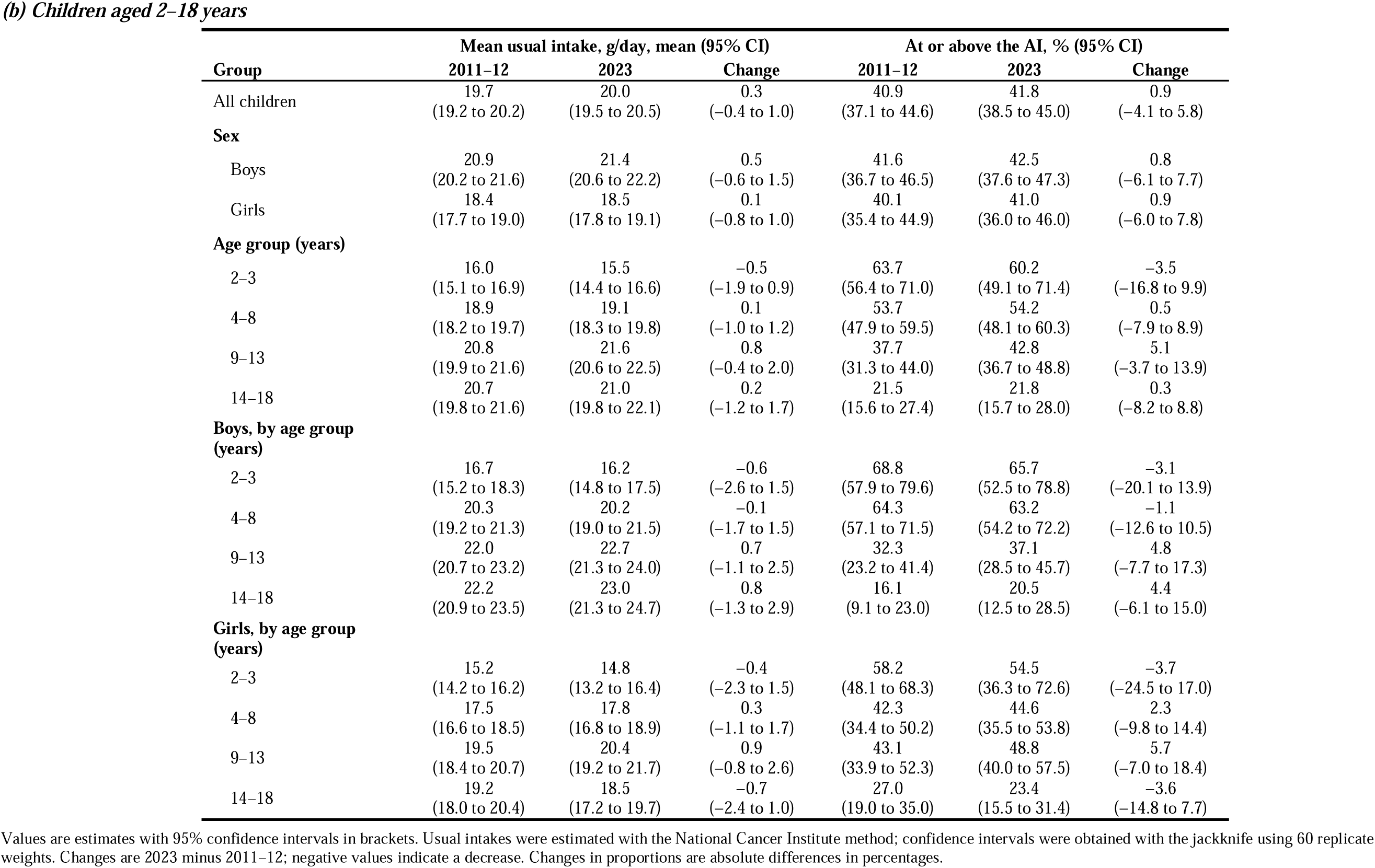
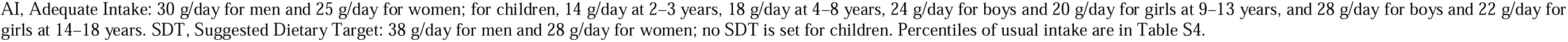
Usual dietary fibre intake in Australian adults and children, National Nutrition and Physical Activity Survey (NNPAS) 2011–12 and 2023.

|  | **Mean usual intake, g/day, mean (95% CI)** |  |  | **At or above the AI, % (95% CI)** |  |  | **At or above the SDT, % (95% CI)** |  |  |
|---|---|---|---|---|---|---|---|---|---|
| **Group** | **2011–12** | **2023** | **Change** | **2011–12** | **2023** | **Change** | **2011–12** | **2023** | **Change** |
| All adults | 22.9<br>(22.5 to 23.3) | 20.8<br>(20.5 to 21.1) | –2.1<br>(–2.6 to –1.6) | 24.6<br>(22.9 to 26.4) | 17.2<br>(15.7 to 18.7) | –7.4<br>(–9.8 to –5.1) | 11.4<br>(9.9 to 12.9) | 7.5<br>(6.4 to 8.6) | –3.9<br>(–5.7 to –2.0) |
| **Sex** |  |  |  |  |  |  |  |  |  |
| Men | 24.5<br>(23.9 to 25.1) | 22.2<br>(21.8 to 22.6) | –2.3<br>(–3.0 to –1.6) | 22.3<br>(20.2 to 24.3) | 15.1<br>(13.4 to 16.8) | –7.2<br>(–9.8 to –4.5) | 6.1<br>(4.7 to 7.4) | 3.7<br>(2.8 to 4.7) | –2.3<br>(–4.0 to –0.7) |
| Women | 21.3<br>(20.8 to 21.9) | 19.5<br>(19.2 to 19.9) | –1.8<br>(–2.4 to –1.2) | 27.0<br>(24.4 to 29.5) | 19.3<br>(17.3 to 21.3) | –7.7<br>(–10.9 to –4.5) | 16.5<br>(14.3 to 18.7) | 11.1<br>(9.4 to 12.7) | –5.5<br>(–8.2 to –2.7) |
| **Age group (years)** |  |  |  |  |  |  |  |  |  |
| 19–30 | 22.2<br>(21.5 to 22.9) | 20.5<br>(19.9 to 21.2) | –1.7<br>(–2.6 to –0.7) | 21.4<br>(18.7 to 24.1) | 16.5<br>(13.6 to 19.4) | –4.9<br>(–8.9 to –1.0) | 9.4<br>(7.4 to 11.5) | 7.4<br>(5.5 to 9.4) | –2.0<br>(–4.8 to 0.8) |
| 31–50 | 22.8<br>(22.3 to 23.2) | 20.9<br>(20.5 to 21.3) | –1.9<br>(–2.5 to –1.2) | 24.0<br>(21.9 to 26.1) | 17.5<br>(15.6 to 19.4) | –6.5<br>(–9.3 to –3.6) | 10.7<br>(9.2 to 12.2) | 7.6<br>(6.2 to 9.0) | –3.1<br>(–5.2 to –1.1) |
| 51–70 | 23.4<br>(22.8 to 24.0) | 20.9<br>(20.4 to 21.3) | –2.5<br>(–3.3 to –1.8) | 27.2<br>(24.5 to 30.0) | 17.4<br>(15.4 to 19.4) | –9.8<br>(–13.3 to –6.4) | 13.2<br>(11.2 to 15.2) | 7.5<br>(6.1 to 8.8) | –5.7<br>(–8.2 to –3.3) |
| 71 and over | 23.3<br>(22.5 to 24.1) | 21.0<br>(20.4 to 21.6) | –2.3<br>(–3.4 to –1.3) | 27.0<br>(22.7 to 31.2) | 17.2<br>(14.7 to 19.7) | –9.8<br>(–14.7 to –4.8) | 13.0<br>(10.2 to 15.8) | 7.4<br>(5.4 to 9.4) | –5.6<br>(–9.0 to –2.2) |
| **Men, by age group (years)** |  |  |  |  |  |  |  |  |  |
| 19–30 | 23.8<br>(22.9 to 24.7) | 21.7<br>(20.8 to 22.7) | –2.1<br>(–3.4 to –0.8) | 19.6<br>(16.2 to 23.0) | 14.0<br>(11.1 to 17.0) | –5.6<br>(–10.1 to –1.1) | 5.0<br>(3.1 to 7.0) | 3.5<br>(2.1 to 4.8) | –1.6<br>(–3.9 to 0.8) |
| 31–50 | 24.5<br>(23.9 to 25.1) | 22.2<br>(21.6 to 22.8) | –2.3<br>(–3.1 to –1.4) | 22.1<br>(19.5 to 24.6) | 15.2<br>(12.9 to 17.6) | –6.8<br>(–10.3 to –3.3) | 5.7<br>(4.3 to 7.0) | 3.8<br>(2.1 to 5.6) | –1.8<br>(–4.0 to 0.3) |
| 51–70 | 24.8<br>(23.9 to 25.7) | 22.4<br>(21.8 to 22.9) | –2.4<br>(–3.4 to –1.4) | 23.8<br>(20.2 to 27.4) | 15.6<br>(13.5 to 17.8) | –8.2<br>(–12.4 to –4.0) | 7.0<br>(4.9 to 9.1) | 3.9<br>(2.7 to 5.2) | –3.1<br>(–5.5 to –0.6) |
| 71 and over | 25.2<br>(24.2 to 26.3) | 22.6<br>(21.8 to 23.3) | –2.7<br>(–4.0 to –1.4) | 25.1<br>(20.7 to 29.4) | 15.2<br>(11.7 to 18.7) | –9.9<br>(–15.5 to –4.3) | 7.5<br>(5.5 to 9.6) | 3.5<br>(1.8 to 5.2) | –4.0<br>(–6.7 to –1.4) |
| **Women, by age group (years)** |  |  |  |  |  |  |  |  |  |
| 19–30 | 20.5<br>(19.7 to 21.3) | 19.3<br>(18.7 to 20.0) | –1.2<br>(–2.2 to –0.1) | 23.3<br>(19.6 to 26.9) | 18.9<br>(15.2 to 22.6) | –4.4<br>(–9.6 to 0.8) | 14.0<br>(10.7 to 17.3) | 11.3<br>(8.3 to 14.4) | –2.6<br>(–7.1 to 1.9) |
| 31–50 | 21.1<br>(20.5 to 21.7) | 19.7<br>(19.2 to 20.2) | –1.4<br>(–2.2 to –0.7) | 25.9<br>(23.1 to 28.8) | 19.8<br>(17.0 to 22.6) | –6.1<br>(–10.2 to –2.1) | 15.7<br>(13.3 to 18.0) | 11.2<br>(9.0 to 13.3) | –4.5<br>(–7.7 to –1.3) |
| 51–70 | 22.1<br>(21.5 to 22.8) | 19.5<br>(18.9 to 20.1) | –2.6<br>(–3.5 to –1.7) | 30.6<br>(27.4 to 33.8) | 19.1<br>(15.8 to 22.3) | –11.5<br>(–16.1 to –6.9) | 19.2<br>(16.4 to 22.0) | 10.8<br>(8.9 to 12.7) | –8.4<br>(–11.8 to –5.0) |
| 71 and over | 21.7<br>(20.8 to 22.6) | 19.6<br>(18.8 to 20.4) | –2.1<br>(–3.3 to –0.9) | 28.5<br>(22.9 to 34.1) | 19.0<br>(15.4 to 22.5) | –9.6<br>(–16.2 to –2.9) | 17.5<br>(13.4 to 21.7) | 10.8<br>(7.8 to 13.9) | –6.7<br>(–11.8 to –1.5) |
|  | **Mean usual intake, g/day, mean (95% CI)** |  |  | **At or above the AI, % (95% CI)** |  |  |  |  |  |
| **Group** | **2011–12** | **2023** | **Change** | **2011–12** | **2023** | **Change** |  |  |  |
| All children | 19.7<br>(19.2 to 20.2) | 20.0<br>(19.5 to 20.5) | 0.3<br>(–0.4 to 1.0) | 40.9<br>(37.1 to 44.6) | 41.8<br>(38.5 to 45.0) | 0.9<br>(–4.1 to 5.8) |  |  |  |
| **Sex** |  |  |  |  |  |  |  |  |  |
| Boys | 20.9<br>(20.2 to 21.6) | 21.4<br>(20.6 to 22.2) | 0.5<br>(–0.6 to 1.5) | 41.6<br>(36.7 to 46.5) | 42.5<br>(37.6 to 47.3) | 0.8<br>(–6.1 to 7.7) |  |  |  |
| Girls | 18.4<br>(17.7 to 19.0) | 18.5<br>(17.8 to 19.1) | 0.1<br>(–0.8 to 1.0) | 40.1<br>(35.4 to 44.9) | 41.0<br>(36.0 to 46.0) | 0.9<br>(–6.0 to 7.8) |  |  |  |
| **Age group (years)** |  |  |  |  |  |  |  |  |  |
| 2–3 | 16.0<br>(15.1 to 16.9) | 15.5<br>(14.4 to 16.6) | –0.5<br>(–1.9 to 0.9) | 63.7<br>(56.4 to 71.0) | 60.2<br>(49.1 to 71.4) | –3.5<br>(–16.8 to 9.9) |  |  |  |
| 4–8 | 18.9<br>(18.2 to 19.7) | 19.1<br>(18.3 to 19.8) | 0.1<br>(–1.0 to 1.2) | 53.7<br>(47.9 to 59.5) | 54.2<br>(48.1 to 60.3) | 0.5<br>(–7.9 to 8.9) |  |  |  |
| 9–13 | 20.8<br>(19.9 to 21.6) | 21.6<br>(20.6 to 22.5) | 0.8<br>(–0.4 to 2.0) | 37.7<br>(31.3 to 44.0) | 42.8<br>(36.7 to 48.8) | 5.1<br>(–3.7 to 13.9) |  |  |  |
| 14–18 | 20.7<br>(19.8 to 21.6) | 21.0<br>(19.8 to 22.1) | 0.2<br>(–1.2 to 1.7) | 21.5<br>(15.6 to 27.4) | 21.8<br>(15.7 to 28.0) | 0.3<br>(–8.2 to 8.8) |  |  |  |
| **Boys, by age group (years)** |  |  |  |  |  |  |  |  |  |
| 2–3 | 16.7<br>(15.2 to 18.3) | 16.2<br>(14.8 to 17.5) | –0.6<br>(–2.6 to 1.5) | 68.8<br>(57.9 to 79.6) | 65.7<br>(52.5 to 78.8) | –3.1<br>(–20.1 to 13.9) |  |  |  |
| 4–8 | 20.3<br>(19.2 to 21.3) | 20.2<br>(19.0 to 21.5) | –0.1<br>(–1.7 to 1.5) | 64.3<br>(57.1 to 71.5) | 63.2<br>(54.2 to 72.2) | –1.1<br>(–12.6 to 10.5) |  |  |  |
| 9–13 | 22.0<br>(20.7 to 23.2) | 22.7<br>(21.3 to 24.0) | 0.7<br>(–1.1 to 2.5) | 32.3<br>(23.2 to 41.4) | 37.1<br>(28.5 to 45.7) | 4.8<br>(–7.7 to 17.3) |  |  |  |
| 14–18 | 22.2<br>(20.9 to 23.5) | 23.0<br>(21.3 to 24.7) | 0.8<br>(–1.3 to 2.9) | 16.1<br>(9.1 to 23.0) | 20.5<br>(12.5 to 28.5) | 4.4<br>(–6.1 to 15.0) |  |  |  |
| **Girls, by age group (years)** |  |  |  |  |  |  |  |  |  |
| 2–3 | 15.2<br>(14.2 to 16.2) | 14.8<br>(13.2 to 16.4) | –0.4<br>(–2.3 to 1.5) | 58.2<br>(48.1 to 68.3) | 54.5<br>(36.3 to 72.6) | –3.7<br>(–24.5 to 17.0) |  |  |  |
| 4–8 | 17.5<br>(16.6 to 18.5) | 17.8<br>(16.8 to 18.9) | 0.3<br>(–1.1 to 1.7) | 42.3<br>(34.4 to 50.2) | 44.6<br>(35.5 to 53.8) | 2.3<br>(–9.8 to 14.4) |  |  |  |
| 9–13 | 19.5<br>(18.4 to 20.7) | 20.4<br>(19.2 to 21.7) | 0.9<br>(–0.8 to 2.6) | 43.1<br>(33.9 to 52.3) | 48.8<br>(40.0 to 57.5) | 5.7<br>(–7.0 to 18.4) |  |  |  |
| 14–18 | 19.2<br>(18.0 to 20.4) | 18.5<br>(17.2 to 19.7) | –0.7<br>(–2.4 to 1.0) | 27.0<br>(19.0 to 35.0) | 23.4<br>(15.5 to 31.4) | –3.6<br>(–14.8 to 7.7) |  |  |  |

Values are estimates with 95% confidence intervals in brackets. Usual intakes were estimated with the National Cancer Institute method; confidence intervals were obtained with the jackknife using 60 replicate weights. Changes are 2023 minus 2011–12; negative values indicate a decrease. Changes in proportions are absolute differences in percentages.
AI, Adequate Intake: 30 g/day for men and 25 g/day for women; for children, 14 g/day at 2–3 years, 18 g/day at 4–8 years, 24 g/day for boys and 20 g/day for girls at 9–13 years, and 28 g/day for boys and 22 g/day for girls at 14–18 years. SDT, Suggested Dietary Target: 38 g/day for men and 28 g/day for women; no SDT is set for children. Percentiles of usual intake are in Table S4.

In children, mean usual intake was 20.0 g/day (95% CI = 19.5 to 20.5), and 41.8% (95% CI = 38.5 to 45.0) met the AI. Attainment fell steeply with age (Figure 2). It was 60.2% at 2–3 years, 54.2% at 4–8 years, 42.8% at 9–13 years and 21.8% at 14–18 years. Adolescents had the lowest attainment of any group in 2023, with 20.5% of boys and 23.4% of girls aged 14–18 years meeting the AI. The AI almost doubles across childhood, from 14 g/day at 2–3 years to 28 g/day for boys and 22 g/day for girls at adolescence, whereas mean usual intake rose only from 15.5 to 21.0 g/day.

**Figure 2.**
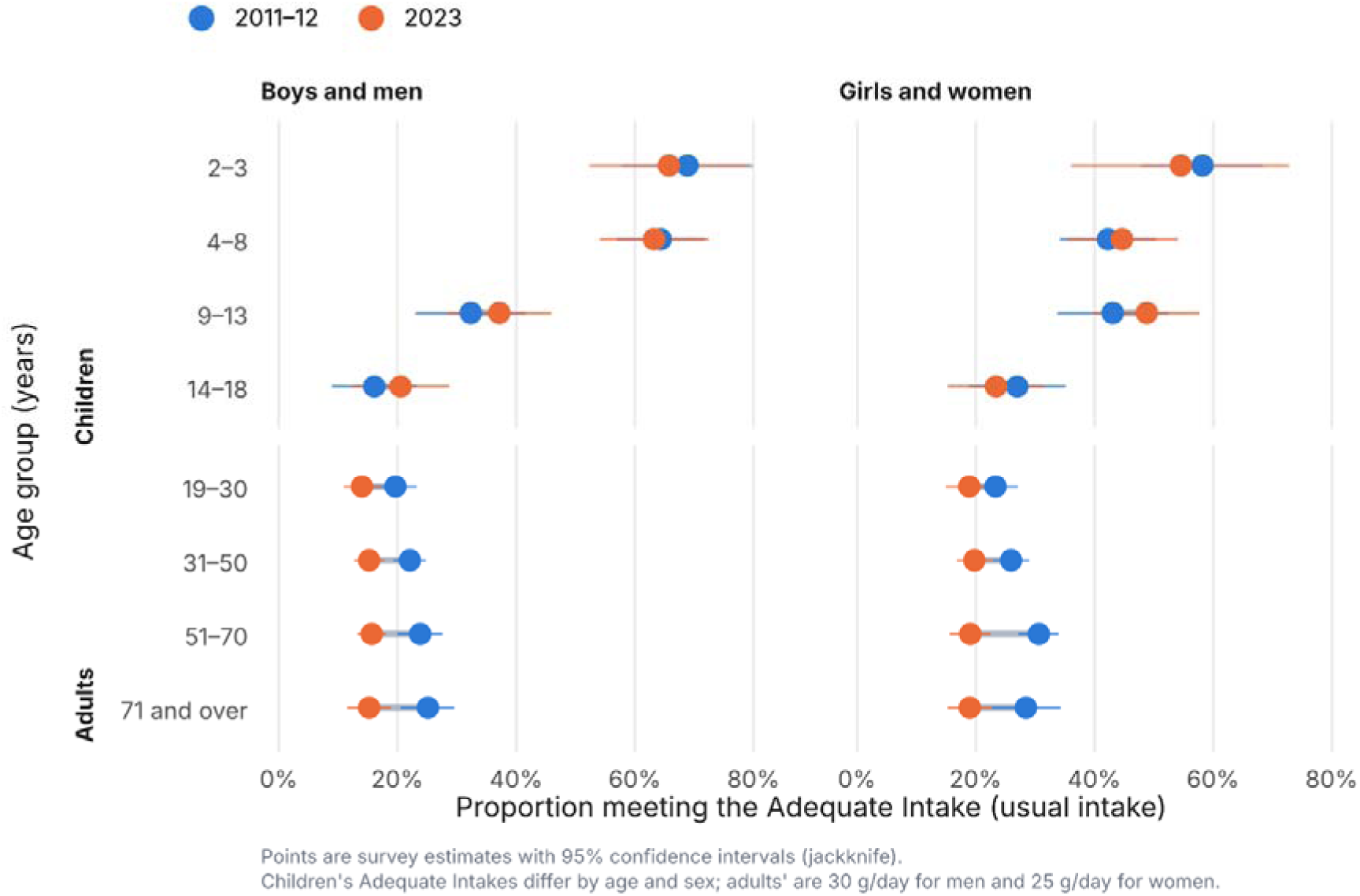
Proportion of children and adults with usual dietary fibre intake at or above the Adequate Intake, by age group and sex, NNPAS 2011–12 and 2023. ^a^ Points are survey estimates with 95% confidence intervals. Children’s Adequate Intakes differ by age and sex; adults’ are 30 g/day for men and 25 g/day for women.

### 3.4 Changes from 2011–12 to 2023

Usual fibre intake in adults fell between the surveys. Mean intake decreased by 2.1 g/day (95% CI = 1.6 to 2.6), from 22.9 to 20.8 g/day, a relative fall of 9.0% (Table 2). The whole distribution shifted to lower intakes, with the median falling from 21.9 to 19.9 g/day (Figure 1; Table S4). Every percentile fell, and each change interval excluded zero: the 5^th^ percentile decreased by 1.5 g/day (95% CI = 0.7 to 2.3) and the 95^th^ by 2.8 g/day (95% CI = 1.4 to 4.2), so the decline was not confined to the top of the distribution. The proportion meeting the AI declined by 7.4% (95% CI = 5.1 to 9.8), and the proportion meeting the SDT declined by 3.9% (95% CI = 2.0 to 5.7). The decline was seen in both men (2.3 g/day, 95% CI = 1.6 to 3.0) and women (1.8 g/day, 95% CI = 1.2 to 2.4).

The decline was larger in older adults. Mean intake dropped by 2.5 g/day (95% CI = 1.8 to 3.3) in adults aged 51–70 years and by 2.3 g/day (95% CI = 1.3 to 3.4) in those aged 71 years and over. AI attainment decreased by 9.8% in both older groups (51–70 years, 95% CI = 6.4 to 13.3; 71 years and over, 95% CI = 4.8 to 14.7). The largest decrease was seen in women aged 51–70 years, whose AI attainment decreased by 11.5% (95% CI = 6.9 to 16.1) and SDT attainment by 8.4% (95% CI = 5.0 to 11.8). As a result, the higher attainment seen in older adults in 2011–12 was no longer present in 2023 (Figure 2).

Usual fibre intake in children did not change. Mean intake was 19.7 g/day in 2011–12 and 20.0 g/day in 2023 (change 0.3 g/day, 95% CI = −0.4 to 1.0). AI attainment was 40.9% and 41.8% (change 0.9%, 95% CI = −4.1 to 5.8). No age or sex group showed a change with a 95% CI that excluded zero.

### 3.5 Subgroup analysis

In 2023, usual fibre intake followed a socioeconomic gradient (Table 3). Adults in the most disadvantaged quintile had a mean intake of 19.5 g/day, compared with 22.1 g/day in the least disadvantaged. AI attainment was 13.9% and 21.4%, respectively. Intake declined in every quintile between the surveys, with reductions ranging from 1.7 g/day (95% CI = 0.6 to 2.8) in quintile 2 to 2.5 g/day (95% CI = 1.4 to 3.6) in quintile 3 (Figure 3).

**Figure 3.**
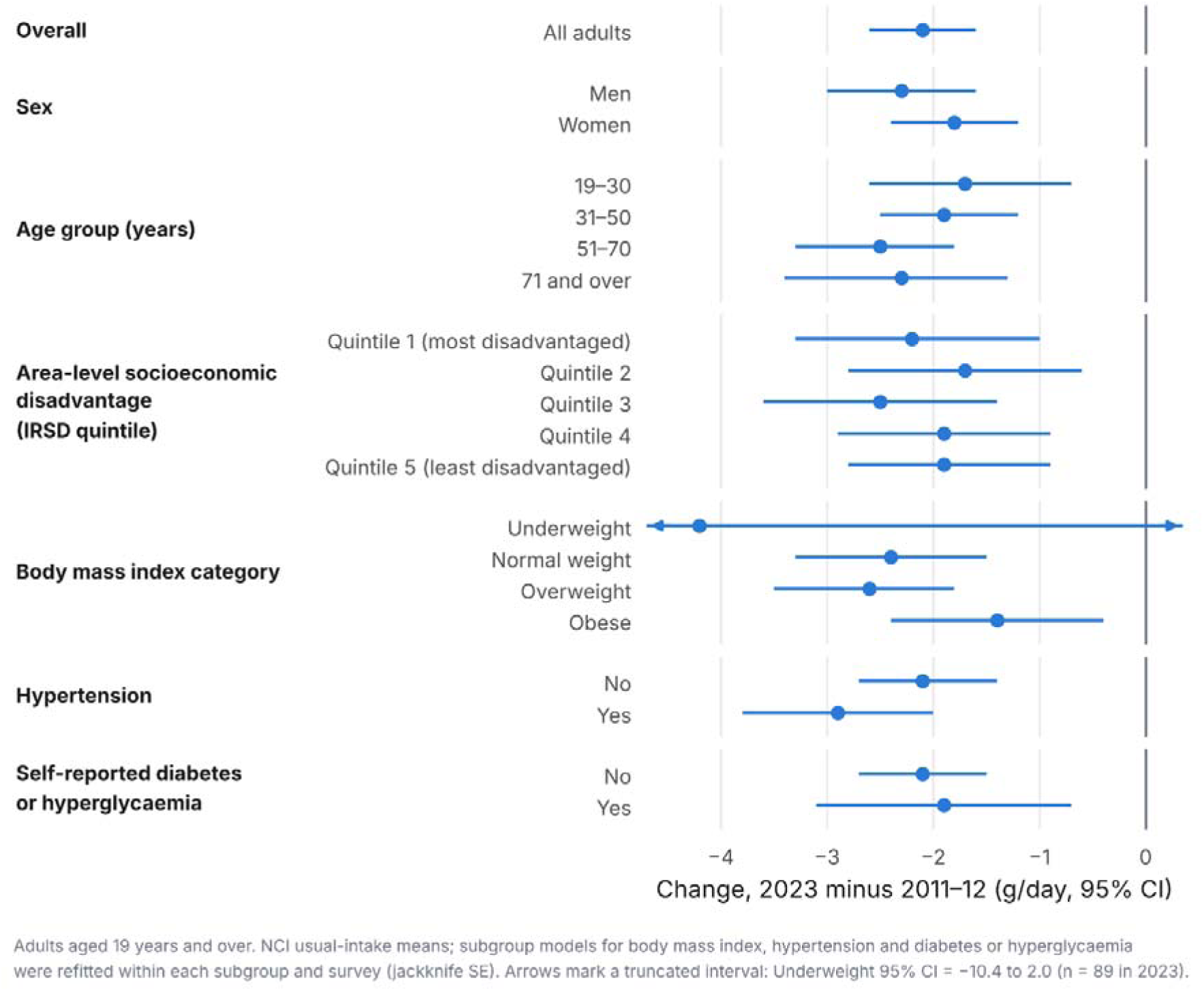
Change in mean usual dietary fibre intake in adults from 2011–12 to 2023, by sociodemographic and health characteristics. Changes are 2023 minus 2011–12, with 95% confidence intervals. ^a^ Estimates by sex, age group and area-level socioeconomic disadvantage come from the main model; estimates by body mass index category, hypertension, and self-reported diabetes or hyperglycemia come from models refit within each group and survey. ^b^ The interval for underweight is truncated (n=89 in 2023). ^c^ IRSD, Index of Relative Socio-economic Disadvantage.

**Table 3.** Usual dietary fibre intake in adults in 2023 by socioeconomic and health characteristics, and change since 2011–12.

| Characteristic | 2023, n | Mean usual intake, g/day, mean (95% CI) |  | At or above the AI, % (95% CI) |  | At or above the SDT, % (95% CI) |  |
|---|---|---|---|---|---|---|---|
|  |  | 2023 | Change | 2023 | Change | 2023 | Change |
| **Area-level socioeconomic disadvantage (IRSD quintile)** |  |  |  |  |  |  |  |
| Quintile 1 (most disadvantaged) | 1,610 | 19.5<br>(18.8 to 20.2) | −2.2<br>(−3.3 to −1.0) | 13.9<br>(10.7 to 17.0) | −5.9<br>(−11.3 to −0.6) | 6.1<br>(3.9 to 8.2) | −2.5<br>(−6.4 to 1.3) |
| Quintile 2 | 1,736 | 20.5<br>(19.8 to 21.2) | −1.7<br>(−2.8 to −0.6) | 15.9<br>(13.0 to 18.8) | −5.5<br>(−9.9 to −1.1) | 6.8<br>(5.0 to 8.5) | −2.7<br>(−5.5 to 0.1) |
| Quintile 3 | 1,800 | 20.4<br>(19.7 to 21.1) | −2.5<br>(−3.6 to −1.4) | 15.5<br>(13.0 to 18.0) | −9.4<br>(−14.0 to −4.8) | 6.6<br>(4.5 to 8.7) | −4.7<br>(−7.9 to −1.5) |
| Quintile 4 | 1,688 | 21.5<br>(20.8 to 22.2) | −1.9<br>(−2.9 to −0.9) | 19.4<br>(16.4 to 22.3) | −7.4<br>(−11.9 to −2.8) | 8.5<br>(6.6 to 10.4) | −3.9<br>(−6.9 to −0.9) |
| Quintile 5 (least disadvantaged) | 1,912 | 22.1<br>(21.6 to 22.6) | −1.9<br>(−2.8 to −0.9) | 21.4<br>(18.9 to 23.8) | −8.1<br>(−12.5 to −3.7) | 9.6<br>(7.9 to 11.2) | −5.0<br>(−8.3 to −1.8) |
| **Body mass index category** |  |  |  |  |  |  |  |
| Underweight | 89 | 19.3<br>(16.5 to 22.0) | −4.2<br>(−10.4 to 2.0) | 16.7<br>(4.2 to 29.2) | −9.0<br>(−43.5 to 25.4) | 8.4<br>(−0.5 to 17.4) | −2.7<br>(−26.6 to 21.2) |
| Normal weight | 2,492 | 21.8<br>(21.2 to 22.4) | −2.4<br>(−3.3 to −1.5) | 22.3<br>(19.3 to 25.3) | −9.2<br>(−13.6 to −4.8) | 10.7<br>(8.5 to 13.0) | −5.7<br>(−9.4 to −2.0) |
| Overweight | 2,991 | 21.1<br>(20.5 to 21.7) | −2.6<br>(−3.5 to −1.8) | 17.7<br>(14.9 to 20.5) | −8.0<br>(−12.0 to −3.9) | 7.5<br>(5.7 to 9.4) | −3.1<br>(−6.0 to −0.1) |
| Obese | 3,032 | 19.7<br>(19.1 to 20.3) | −1.4<br>(−2.4 to −0.4) | 11.1<br>(8.3 to 13.8) | −4.3<br>(−8.8 to 0.2) | 3.8<br>(2.1 to 5.4) | −1.9<br>(−4.6 to 0.8) |
| **Self-reported hypertension** |  |  |  |  |  |  |  |
| No | 5,503 | 21.2<br>(20.7 to 21.6) | −2.1<br>(−2.7 to −1.4) | 18.7<br>(16.3 to 21.2) | −7.0<br>(−10.5 to −3.4) | 8.3<br>(6.6 to 9.9) | −3.4<br>(−6.0 to −0.9) |
| Yes | 3,169 | 20.0<br>(19.5 to 20.5) | −2.9<br>(−3.8 to −2.0) | 13.6<br>(11.2 to 16.0) | −10.6<br>(−14.9 to −6.3) | 5.4<br>(3.8 to 7.0) | −5.6<br>(−8.8 to −2.3) |
| **Self-reported diabetes or hyperglycaemia** |  |  |  |  |  |  |  |
| No | 7,478 | 20.8<br>(20.4 to 21.2) | −2.1<br>(−2.7 to −1.5) | 16.8<br>(15.2 to 18.5) | −7.7<br>(−10.6 to −4.8) | 7.2<br>(6.1 to 8.3) | −4.1<br>(−6.3 to −1.9) |
| Yes | 1,268 | 21.3<br>(20.4 to 22.2) | −1.9<br>(−3.1 to −0.7) | 19.6<br>(14.6 to 24.6) | −6.3<br>(−12.8 to 0.3) | 9.2<br>(5.7 to 12.6) | −3.0<br>(−7.9 to 1.8) |

Adults aged 19 years and older. Values are estimates with 95% confidence intervals in brackets. Usual intakes were estimated with the National Cancer Institute method; confidence intervals were obtained with the jackknife using 60 replicate weights. Changes are 2023 minus 2011–12; negative values indicate a decrease. Changes in proportions are absolute differences in percentages. n is the unweighted number of adults in 2023.
Estimates by area-level socioeconomic disadvantage come from the main usual-intake model; estimates by body mass index category, hypertension, and self-reported diabetes or hyperglycaemia come from models refitted within each group and survey. Definitions are given in Section 2.2 and Table S1.
AI, Adequate Intake (30 g/day for men and 25 g/day for women); IRSD, Index of Relative Socio-economic Disadvantage; SDT, Suggested Dietary Target (38 g/day for men and 28 g/day for women).

In 2023, adults with obesity had the lowest fibre intake (19.7 g/day), and 3.8% met the SDT, compared with 10.7% of those with normal weight (Figure 3). Since 2011–12, intake fell by 2.4 g/day (95% CI = 1.5 to 3.3) in adults with normal weight, 2.6 g/day (95% CI = 1.8 to 3.5) in those with overweight and 1.4 g/day (95% CI = 0.4 to 2.4) in those with obesity. SDT attainment decreased by 5.7% (95% CI = 2.0 to 9.4), 3.1% (95% CI = 0.1 to 6.0) and 1.9% (95% CI = −0.8 to 4.6), respectively.

Adults with hypertension had a lower fibre intake than those without (20.0 versus 21.2 g/day), and fewer met the SDT (5.4% versus 8.3%). They also had a larger decline since 2011–12, both in mean intake (a reduction of 2.9 g/day, 95% CI = 2.0 to 3.8, versus 2.1 g/day, 95% CI = 1.4 to 2.7) and in SDT attainment (a drop of 5.6%, 95% CI = 2.3 to 8.8, versus 3.4%, 95% CI = 0.9 to 6.0).

Intake in adults with self-reported diabetes or hyperglycaemia was similar to that in adults without (21.3 versus 20.8 g/day). SDT attainment was also similar (9.2% versus 7.2%), as were the declines in intake over time (1.9 g/day, 95% CI = 0.7 to 3.1, versus 2.1 g/day, 95% CI = 1.5 to 2.7).

### 3.6 Usual fibre density

In 2023, mean usual fibre density was 10.7 g/1,000 kcal (95% CI = 10.5 to 10.8) in adults and 10.4 g/1,000 kcal (95% CI = 10.2 to 10.6) in children. Only 13.9% of adults (95% CI = 12.2 to 15.6) and 7.3% of children (95% CI = 3.6 to 10.9) reached the benchmark of 14 g/1,000 kcal. Since 2011–12, fibre density in adults fell less than fibre intake (3.2% versus 9.0%; Figure 4), so most of the decline in fibre accompanied lower energy intake. The proportion of adults reaching the benchmark decreased by 4.0% (95% CI = 1.4 to 6.6), mainly among women (from 24.2% to 17.7%), whose mean density also fell by 0.66 g/1,000 kcal (95% CI = 0.33 to 0.98). Men and children showed no clear change.

**Figure 4.**
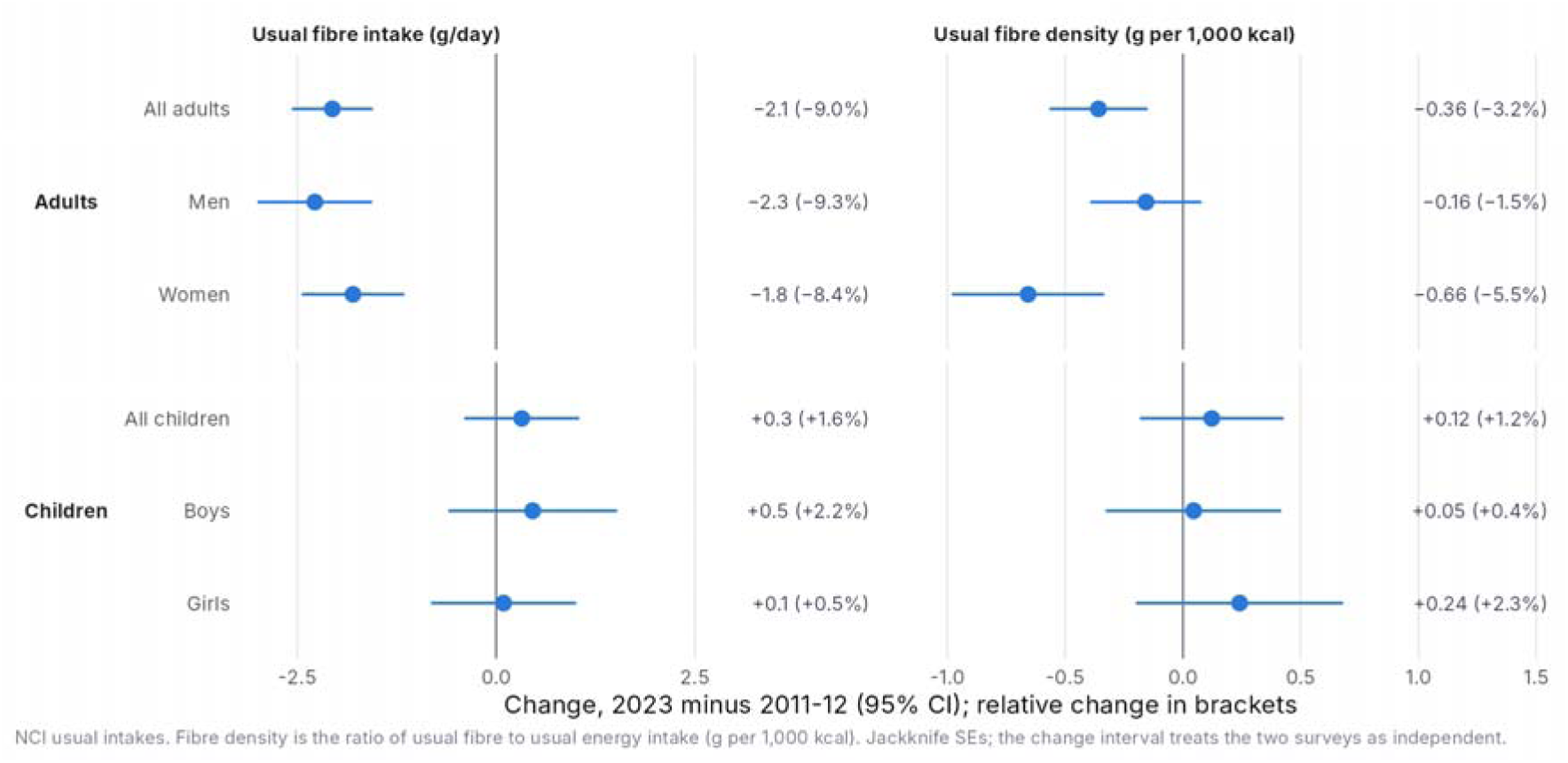

## 4. Discussion

Using repeat 24-hour recalls from two nationally representative surveys, this study found that usual dietary fibre intake among Australian adults declined by approximately 2 g/day over the last decade. By 2023, fewer than one in five adults met the AI, and only about one in 13 met the SDT. Declines occurred in both sexes and across adult age groups, with some of the largest reductions among adults aged 51 years and over. Fibre intake among children remained stable overall, with 41.8% reaching AI, although AI attainment decreased markedly with age and was lowest among adolescents.

The lower adult intake in 2023 contrasts with the relative stability seen in earlier Australian surveys. Mean adult fibre intake was 23.1 g/day in the 1995 National Nutrition Survey [30], and an analysis of the 2011–12 NNPAS concluded that intake had changed little over the preceding 10–20 years [7]. Our usual-intake estimate of 22.9 g/day for 2011–12 is consistent with these earlier findings. Although dietary assessments differed across these three surveys, the 2023 estimate is the first national estimate since 1995 to show a clear decrease in adult fibre intake. Other population data are consistent with changes in the consumption of some fibre-rich foods: the ABS reported lower median consumption of fruit products and dishes among consumers in 2023 than in 2011–12 [31].

Trends in fibre intake differ across countries. The United Kingdom and the Netherlands reported increases in adult fibre intake across national surveys over the last decade [32,33]. In the United States, mean fibre intake remained stable at about 16 g/day between 1999 and 2008 [34], and rose only slightly to 2017, with a fall among non-Hispanic White adults [35]. By contrast, intake has declined in several Asian countries. In Japan, fibre intake fell by about 30% between 1952 and 1970 and changed little thereafter [36]. In China, grain and fibre intakes have fallen continuously since 1982, as consumption of refined grains increased [37]. In Korea, intakes of fruit, vegetables, whole grains and fibre declined between 2013 and 2022 [38]. Therefore, the declines in Australia resembles the trends in several Asian countries. However, these comparisons should be interpreted with caution. Population data for most countries are not recent, and countries differ in their fibre recommendations and dietary assessment methods, all of which could contribute to the differences observed.

The decline in adult fibre intake was not explained entirely by lower reported food and energy intake. Fibre density also fell, so each unit of energy in 2023 came from foods containing less fibre, rather than from smaller amounts of the same diet. This raises the question of why intake fell despite long-term recommendations to eat fibre-rich foods [23]. This pattern reflects potential changes in both the food supply and eating habits. Ultra-processed foods are one example. They supplied 42% of energy in the 2011–12 NNPAS, and a higher share was associated with lower fibre intake [39]. Studies in other countries reached similar conclusions [40,41]. Eating away from home, including takeaway foods, has also become more common and is associated with higher energy and fat intake and lower micronutrient intake [42]. In Australia, consumption data show rising purchases of potato chips, chocolate and cereal-based mixed dishes such as pizza and pasta, alongside falling purchases of fruit and bread [43]. Identifying which foods drove the decline is therefore a priority, and future food-source analyses of both surveys are needed to guide targeted strategies.

Despite the decline, mean fibre intake among Australian adults in 2023 remained similar to or higher than estimates from several comparable countries. Reported adult intakes are approximately 15–17 g/day in the United States and United Kingdom, 16–18 g/day in Canada, 18–22 g/day in New Zealand and 18–20 g/day in European populations [3–6,9,44]. Direct comparisons are limited by differences in survey periods, dietary assessment methods, population age ranges and national reference values. Nevertheless, the consistently low proportions reaching country-specific recommendations, particularly comparable high-income countries, indicate that low fibre intake is a common population-health concern rather than a problem unique to Australia.

Age-related findings identified two groups of particular concern. Adults aged 51 years and over, who had some of the highest intakes in 2011–12, experienced some of the largest declines by 2023. Fibre intake among children remained stable overall, but AI attainment was lower at older ages, from 60.2% at 2–3 years to 21.8% at 14–18 years. This reflects an increase in age-specific reference values that was not matched by a comparable increase in intake. Low whole-grain consumption may contribute to the adolescent shortfall [45,46], while the decline among older adults is concerning given their increasing risk of cardiovascular disease and type 2 diabetes [1]. Together, these findings identify adolescence and later adulthood as potentially important life stages for intervention.

Lower fibre intake was also observed among adults living in socioeconomically disadvantaged areas and those with cardiometabolic risk factors. In 2023, adults living in the most disadvantaged areas consumed approximately 2.6 g/day less fibre than those in the least disadvantaged areas, consistent with earlier Australian evidence of socioeconomic differences in fibre intake and overall diet quality [7,47]. Differences in the affordability and accessibility of fibre-rich foods may contribute to this gradient [48,49]. In subgroup analyses, intake was also lower among adults with hypertension and obesity. These differences are clinically relevant because higher fibre intake is associated with improved cardiovascular and metabolic outcomes [4,50]. However, differential dietary under-reporting may partly explain the findings for obesity [51,52]. These subgroup results should, therefore, be interpreted as descriptive rather than causal.

This study has several strengths. It utilised the most recent national data to compare usual fibre intake across two nationally representative Australian surveys, with repeat 24-hour recalls in a large subsample of each. Usual intake was estimated with the NCI method, which removes day-to-day variation, so the proportions meeting the AI and SDT reflect habitual intake rather than intake on a single day [11,12]. Uncertainty was estimated with the ABS replicate weights, and modelling fibre jointly with energy separated changes in diet composition from changes in the amount eaten. However, some limitations should be noted. Dietary assessment methods and food composition databases differed between the surveys, and self-reported intake is prone to under-reporting. The available microdata lacked detailed health information, which limited further subgroup analyses, such as by diabetes type or cardiovascular disease. Finally, food sources of fibre were not analysed, so the foods that drove the decline could not be identified.

In conclusion, usual fibre intake among Australian adults was lower in 2023 than in 2011–12, with fewer than one in five having an intake at or above the AI and only one in thirteen meeting SDT for prevention of non-communicable chronic disease. The decline reflects both lower energy intake and less fibre-rich food choices. Children’s intake remained stable overall, but AI attainment was particularly low among adolescents. Older adults had the largest declines, and intake was lower among socioeconomically disadvantaged adults and those with hypertension or obesity. Public health strategies that improve the affordability, availability and uptake of fibre-rich foods may help narrow the gap between current intakes and dietary targets. Continued surveillance using consistent dietary assessment methods will be important to determine whether the observed decline and population differences persist.

## Data availability

The 2011–12 and 2023 NNPAS Basic Microdata are available to approved users from the Australian Bureau of Statistics.

## Data Availability

All data produced are available online at https://www.abs.gov.au/statistics/health/food-and-nutrition/national-nutrition-and-physical-activity-survey/latest-release

## Supplementary materials

**Table S1.** Harmonisation of variables between the NNPAS 2011–12 and 2023.

**Table S2.** STROBE-nut checklist.

**Table S3.** Dietary fibre intake from a single Day 1 recall and as usual intake, NNPAS 2011–12 and 2023.

**Table S4.** Percentiles of usual dietary fibre intake, NNPAS 2011–12 and 2023.

**Figure S1.** Flow of participants in the NNPAS 2011–12 and 2023.

**Figure S2.** Agreement between Day 1 and Day 2 fibre intake quartiles in adults aged 19 years and over with two recalls, NNPAS 2011–12 and 2023.

## Abbreviations

ABS: Australian Bureau of Statistics
AI: Adequate Intake
AMPM: Automated Multiple-Pass Method
AUSNUT: Australian Food and Nutrient Database
BMI: body mass index
CI: confidence interval
FSANZ: Food Standards Australia New Zealand
IRSD: Index of Relative Socio-economic Disadvantage
MCMC: Markov chain Monte Carlo
NCI: National Cancer Institute
NNPAS: National Nutrition and Physical Activity Survey
SDT: Suggested Dietary Target
SE: standard error
SEIFA: Socio-Economic Indexes for Areas
STROBE-nut: Strengthening the Reporting of Observational Studies in Epidemiology–Nutritional Epidemiology.

