## Supplementary material for "Usual dietary fibre intake in Australia: A comparison of the 2011–12 and 2023 National Nutrition and Physical Activity Surveys": Online supplementary tables and figures

##### Contents

Table S1. Harmonisation of variables between the NNPAS 2011–12 and 2023

Table S2. STROBE-nut checklist

Table S3. Dietary fibre intake from a single Day 1 recall and as usual intake, NNPAS 2011–12 and 2023

Table S4. Percentiles of usual dietary fibre intake, NNPAS 2011–12 and 2023

Figure S1. Flow of participants in the NNPAS 2011–12 and 2023

Figure S2. Agreement between Day 1 and Day 2 fibre intake quartiles in adults, NNPAS 2011–12 and 2023

*NNPAS, National Nutrition and Physical Activity Survey; STROBE-nut, Strengthening the Reporting of Observational Studies in Epidemiology–Nutritional Epidemiology.*

### Supplementary Tables

**Table S1.** Harmonisation of variables between the National Nutrition and Physical Activity Survey (NNPAS) 2011–12 and 2023.

| Variable | ABS item, 2011–12 | ABS item, 2023 | Categories used in the analysis | Comparability notes |
|---|---|---|---|---|
| **Dietary data** | | | | |
| Dietary fibre, Day 1 and Day 2 | FIBREF1 Dietary fibre (food) Day 1 g; FIBREF2 Dietary fibre (food) Day 2 g | FIBREF1 Dietary fibre (food) Day 1 g; FIBREF2 Dietary fibre (food) Day 2 g | g/day | ABS flag: comparable. Different food composition databases: AUSNUT 2011–13 (2011–12) and AUSNUT 2023 (2023). |
| Energy intake, Day 1 | ENERGYF1 Energy with dietary fibre (food) Day 1 kJ | ENERGYF1 Energy with dietary fibre (food) Day 1 kJ | kJ/day, converted to kcal (kJ ÷ 4.184) for fibre density | ABS flag: comparable. Same database difference as fibre. |
| Dietary assessment method | Interviewer-administered Automated Multiple-Pass Method (AMPM) | Intake24; Day 1 interviewer-guided | Not harmonised; described in Section 2.3 | Change in recall instrument and mode between surveys. |
| Second recall completed (Day 2) | NUMRECAL Number of 24hr recalls = 2 and NPAD2WGT > 0 (Person weight, Day 2) | NUMRECAL Number of 24hr recall days = 2 and NPAFWTD2 > 0 (Selected person weight, Day 2) with FIBREF2 not missing | Yes; no | Completion differs: 63.6% of the 2011–12 sample and 80.6% of the 2023 sample. |
| Day of week of recall | NUTDAY1, NUTDAY2 Day of week of day 1 (day 2) consumption: 1 Sunday … 7 Saturday | NUTDAY1, NUTDAY2 Day of the week of consumption (Day 1) (Day 2): 1 Monday … 7 Sunday | Weekend: codes 1 and 7 (2011–12); codes 6 and 7 (2023) | ABS flag: comparable. Coding differs between surveys, so each is recoded separately. |
| **Weights** | | | | |
| Person weight and replicate weights | NPAFINWT Person weight; WPM0101–WPM0160 | NPAFINWT Selected person weight; WPM0101–WPM0160 | Day 1 weight for all analyses | Same structure in both surveys (60 group jackknife replicates). Not recalibrated after the removal of implausible values. |
| **Sociodemographic characteristics** | | | | |
| Age group | AGEC Age of person (continuous values from 2 to 84 years; 85 years and over) | AGEC Age of person (age in single years, 2 to 85) | Children: 2–3, 4–8, 9–13, 14–18 years. Adults: 19–30, 31–50, 51–70, 71 years and over | Same variable. Adults are 19 years and over. |
| Sex | SEX Sex of person: 1 Male; 2 Female | SEX Sex at birth: 1 Male; 2 Female | Male; female | Only codes 1 and 2 occur in both files. |
| Born in Australia | COBBC Country of birth: 1 Australia; 2 Main English Speaking Countries (Canada, Republic of Ireland, New Zealand, South Africa, United Kingdom, United States of America); 3 Other | COBBC Country of birth - SACC code: the same three categories | Born in Australia (code 1); born elsewhere (codes 2–3) | Same coding. |
| Area-level socioeconomic disadvantage (IRSD quintile) | SF2SA1QN Index of Relative Socio-economic Disadvantage - 2011 - SA1 - Quintiles - National: 1 Lowest 20%; 2 Second quintile; 3 Third quintile; 4 Fourth quintile; 5 Highest 20%[a] | SF2SA1QN SEIFA - Index of Relative Socio-economic Disadvantage - 2021 - SA1 - Quintiles - National: 1 First quintile (lowest); 2 Second quintile; 3 Third quintile; 4 Fourth quintile; 5 Fifth quintile (highest)[a] | Quintile 1 (most disadvantaged) to quintile 5 (least disadvantaged) | Based on SEIFA 2011 and SEIFA 2021, so quintiles are relative within each year. |
| Equivalised household income quintile | INCDEC Equivalised income of household: deciles: 01 First decile … 10 Tenth decile; 98 Not stated; 99 Not known; 00 Not applicable | INCDECU1 Equivalised household current weekly income from all sources: deciles: 1 First decile … 10 Tenth decile; 99 Not known/not stated | Pairs of deciles combined into quintiles (the ABS reports deciles only) | ABS flag: comparable. Table 1 only. |
| Remoteness | ARIABC (ASGS 2011): 1 Major cities of Australia; 2 Inner regional Australia; 3 Other | ARIA21BC Remoteness area (ARIA) - ASGS 2021: 1 Major Cities of Australia; 2 Inner Regional Australia; 3 Other | Major cities; inner regional; outer regional and remote | Same three codes in both surveys. |
| Highest non-school qualification (adults) | LVHNSQBC Level of highest non-school qualification: 1 Postgraduate Degree, Graduate Diploma and Graduate Certificate; 2 Bachelor Degree; 3 Advanced Diploma and Diploma; 4 Certificate III and IV; 5 Certificate I and II; 6 Certificate n.f.d.; 7 No non-school qualification; 8 Level not determined | LVHNSQBC: the same eight categories | Bachelor degree or above (codes 1–2); certificate or diploma (codes 3–6); none (code 7). Code 8 set to missing | ABS flag for the full 2023 item (NONSHLVD): comparable. |

| Variable | ABS item, 2011–12 | ABS item, 2023 | Categories used in the analysis | Comparability notes |
|---|---|---|---|---|
| Labour force status (adults) | LFSBC Labour force status: 1 Employed; 2 Unemployed; 3 Not in the labour force | LFS Labour force status: 1 Employed; 2 Unemployed; 3 Not in the labour force | The three ABS categories | ABS flag: new item in 2023, but the categories are the same. |
| Married (adults) | SMSBC Social marital status: 1 Married; 2 Not married | SMSBC Social marital status: 1 Married; 2 Not married | Married; not married | ABS flag: not comparable (derivation change) for the full item SOCMAR. Table 1 only. |
| Living alone (adults) | HHSIZECB Number of persons in household: 1 One (2 Two … 6 Six or more) | HHSIZECB Number of persons in household: 1 One (2 Two … 6 Six or more) | One person (living alone); two or more persons | Table 1 only. |
| **Health characteristics** | | | | |
| Current smoker (adults) | SMKSTAT Smoker status: 1 Current smoker daily; 2 Current smoker weekly (at least once a week but not daily); 3 Current smoker less than weekly; 4 Ex-smoker; 5 Never smoked | SMOKSTAT Current smoker status: 1 Current daily smoker; 2 Current smoker - other; 3 Current non-smoker | Current smoker (2011–12 codes 1–3; 2023 codes 1–2); not current smoker (2011–12 codes 4–5; 2023 code 3) | ABS flag: comparable. |
| Body mass index category | BMICATHY Body Mass Index - measured - categories (Half year scale for children; 18+ for adults): 1 Underweight Class 3; 2 Underweight Class 2; 3 Underweight Class 1; 4 Normal range; 5 Normal range (Adult only); 6 Overweight; 7 Obese Class 1; 8 Obese Class 2 (Adult only); 9 Obese Class 3 (Adult only); 98 Measurement not taken - refusal; 99 Measurement not taken - other reason | BMICATHY Body mass index - measured - categories (half year scale for children, 18+ for adults): categories 1–9 as in 2011–12; 99 Unable to determine | Underweight (codes 1–3); normal weight (codes 4–5); overweight (code 6); obese (codes 7–9) | ABS flag: not comparable (derivation change). In 2011–12, 1,975 people have no category (mostly refusals; pregnant women are coded 99). In 2023, 153 have none (74 pregnant women). 2023 values include some imputed by the ABS (MEASWEIG, MEASHEIG). |
| Hypertension (adults) | Derived from HYPBC Whether has hypertensive disease (1–2 Ever told has hypertensive disease, still current; 3 not current; 5 Never told) and BPCAT Blood pressure categories (18 yrs +) (3 High (from 140/90 to <160/100); 4 Very high; 5 Severe (from 180/110)). Yes = HYPBC 1–2 or BPCAT 3–5; no = HYPBC 3 or 5 and BPCAT 1–2; otherwise missing. Pregnant women (SABDYMS 4 Currently pregnant) set to missing | HHYPTEN Whether has hypertension (self-reported or measured high blood pressure): 1 Has hypertension; 5 Does not have hypertension (adults not pregnant) | Yes; no | ABS flag: HHYPTEN is a new item and BPCAT a derivation change, so definitions are not identical. Pregnant women are excluded in both surveys. Of 8,672 adults in 2023, 2,735 (31.5%) have ABS-imputed blood pressure (BPIMPUTE 2 Measurement imputed). In 2011–12, 1,215 adults have no classification (1,336 had no measured reading, and none were imputed). |
| Self-reported diabetes or hyperglycaemia (adults) | AGDBBC Age first told had diabetes or high sugar levels: 1 Under 25 … 6 65 and over; 97 Not applicable; 99 Not known | AGDBBC Age first told had diabetes or high sugar levels: 1 Under 25 … 6 65 and over; 97 Not applicable; 99 Not known | Yes (codes 1–6 and 99); no (code 97) | Same variable and coding in both surveys. Includes high blood sugar without a diabetes diagnosis, so it does not identify current diabetes. |
| Pregnancy (women) | FEMLSBC Female life stages (single response): 2 Currently pregnant; 3 Currently breastfeeding. SABDYMS 4 Currently pregnant | NHMRCFLS NHMRC reference female lifestage (aged 18–50): 1 Pregnant only; 2 Breastfeeding only; 3 Pregnant and breastfeeding; 4 Neither pregnant nor breastfeeding; 5 Not known or not stated. SABDYMS 4 Currently pregnant | Pregnant women excluded from body mass index categories and hypertension | About 116 pregnant women aged 18–50 years in 2011–12 and 74 in 2023. |

Item names and category labels are given as they appear in the ABS data item lists for the 2011–12 and 2023 Basic Microdata. The ABS comparability flag is taken from the full 2023 data item list. Where an analysis category groups several ABS categories, the ABS codes it combines are listed.

[a] ABS footnote, both surveys: includes records where ARIABC (ARIA21BC) = 3 and SF2SA1QN = 4 or 5.

ABS, Australian Bureau of Statistics; ASGS, Australian Statistical Geography Standard; IRSD, Index of Relative Socio-economic Disadvantage; SA1, Statistical Area Level 1; SEIFA, Socio-Economic Indexes for Areas.

**Table S2.** STROBE-nut checklist: Strengthening the Reporting of Observational Studies in Epidemiology–Nutritional Epidemiology.

| Item no. | Recommendation | Where reported |
|---|---|---|
| **Title and abstract** | | |
| 1 | (a) Indicate the study's design with a commonly used term in the title or the abstract. | Title; Abstract (Methods) |
| nut-1 | State the dietary/nutritional assessment method(s) used in the title, abstract, or keywords. | Abstract (Methods); Keywords |
| | (b) Provide in the abstract an informative and balanced summary of what was done and what was found. | Abstract |
| **Introduction** | | |
| 2 | Background/rationale: explain the scientific background and rationale for the investigation being reported. | Section 1, paragraphs 1–3 |
| 3 | Objectives: state specific objectives, including any prespecified hypotheses. | Section 1, paragraph 4 |
| **Methods** | | |
| 4 | Study design: present key elements of study design early in the paper. | Section 2.1 |
| 5 | Setting: describe the setting, locations, and relevant dates, including periods of recruitment, exposure, follow-up, and data collection. | Section 2.1 |
| nut-5 | Describe any characteristics of the study settings that might affect the dietary intake or nutritional status of the participants, if applicable. | Sections 2.1, 2.3 and 2.5 (survey periods; recall day; weekday and weekend weighting) |
| 6 | (a) Participants (cross-sectional study): give the eligibility criteria and the sources and methods of selection of participants. | Section 2.1 |
| nut-6 | Report particular dietary, physiological, or nutritional characteristics that were considered when selecting the target population. | Sections 2.1 and 2.2 (no selection on diet; pregnant women excluded from BMI and hypertension estimates); Section 2.4 |
| | (b) Matched studies: give matching criteria. | Not applicable |
| 7 | Variables: clearly define all outcomes, exposures, predictors, potential confounders, and effect modifiers. Give diagnostic criteria, if applicable. | Sections 2.2, 2.4 and 2.6; Table S1 |
| nut-7.1 | Clearly define foods, food groups, nutrients, or other food components. | Section 2.4 (definition of dietary fibre) |
| nut-7.2 | When using dietary patterns or indices, describe the methods to obtain them and their nutritional properties. | Not applicable |
| 8 | Data sources/measurement: for each variable of interest, give sources of data and details of methods of assessment. Describe comparability of assessment methods if there is more than one group. | Sections 2.2–2.4; Table S1 |
| nut-8.1 | Describe the dietary assessment method(s), e.g., portion size estimation, number of days and items recorded, how it was developed and administered, and how quality was assured. Report if and how supplement intake was assessed. | Section 2.3; Section 2.4 (supplements) |
| nut-8.2 | Describe and justify food composition data used. Explain the procedure to match food composition with consumption data. Describe the use of conversion factors, if applicable. | Sections 2.3 and 2.4 (AUSNUT 2011–13 and AUSNUT 2023; energy conversion) |
| nut-8.3 | Describe the nutrient requirements, recommendations, or dietary guidelines and the evaluation approach used to compare intake with the dietary reference values, if applicable. | Section 2.4 |
| nut-8.4 | When using nutritional biomarkers, additionally use STROBE-ME. Report the type of biomarkers used and their usefulness as dietary exposure markers. | Not applicable |
| nut-8.5 | Describe the assessment of nondietary data (e.g., nutritional status and influencing factors) and timing of the assessment of these variables in relation to dietary assessment. | Section 2.2 |
| nut-8.6 | Report on the validity of the dietary or nutritional assessment methods and any internal or external validation used in the study, if applicable. | Section 2.3; Section 4 (strengths and limitations) |
| 9 | Bias: describe any efforts to address potential sources of bias. | Sections 2.5 and 2.6 |
| nut-9 | Report how bias in dietary or nutritional assessment was addressed, e.g., misreporting, changes in habits as a result of being measured, or data imputation from other sources. | Section 2.5 (removal of within-person variation; extreme and implausible values); Section 4 (under-reporting) |
| 10 | Study size: explain how the study size was arrived at. | Section 2.1; Section 3.1; Figure S1 |
| 11 | Quantitative variables: explain how quantitative variables were handled in the analyses. If applicable, describe which groupings were chosen and why. | Sections 2.2 and 2.4 |
| nut-11 | Explain the categorization of dietary/nutritional data (e.g., use of N-tiles and handling of nonconsumers) and the choice of reference category, if applicable. | Section 2.4 (reference values; 14 g/1,000 kcal); Section 2.5 (fibre consumed daily) |
| 12 | (a) Describe all statistical methods, including those used to control for confounding. | Sections 2.5 and 2.6 |
| | (b) Describe any methods used to examine subgroups and interactions. | Section 2.6 |
| | (c) Explain how missing data were addressed. | Section 2.2 |

| Item no. | Recommendation | Where reported |
|---|---|---|
| | (d) Cross-sectional study: if applicable, describe analytical methods taking account of sampling strategy. | Sections 2.5 and 2.6 (survey and replicate weights) |
| | (e) Describe any sensitivity analyses. | Not applicable; no sensitivity analyses were conducted |
| nut-12.1 | Describe any statistical method used to combine dietary or nutritional data, if applicable. | Section 2.4 |
| nut-12.2 | Describe and justify the method for energy adjustments, intake modeling, and use of weighting factors, if applicable. | Sections 2.4–2.6 |
| nut-12.3 | Report any adjustments for measurement error, i.e., from a validity or calibration study. | Section 2.5 (National Cancer Institute method; no calibration study) |
| **Results** | | |
| 13 | (a) Report the numbers of individuals at each stage of the study. (b) Give reasons for nonparticipation at each stage. (c) Consider use of a flow diagram. | Sections 2.1 and 3.1; Figure S1 |
| nut-13 | Report the number of individuals excluded based on missing, incomplete, or implausible dietary/nutritional data. | Sections 2.5 and 3.1; Figure S1 |
| 14 | (a) Give characteristics of study participants and information on exposures and potential confounders. (b) Indicate the number of participants with missing data for each variable of interest. | Section 3.1; Table 1 and footnotes |
| nut-14 | Give the distribution of participant characteristics across the exposure variables if applicable. Specify if food consumption of total population or consumers only were used to obtain results. | Table 1; Section 2.5 (total population) |
| 15 | Outcome data (cross-sectional study): report numbers of outcome events or summary measures. | Sections 3.3–3.6; Tables 2 and 3; Figures 1–4 |
| 16 | (a) Give unadjusted estimates and, if applicable, confounder-adjusted estimates and their precision (e.g., 95% confidence interval). (b) Report category boundaries when continuous variables were categorized. (c) If relevant, translate relative risk into absolute risk. | Sections 3.3–3.6; Tables 2 and 3 (descriptive estimates with 95% CIs); Sections 2.2 and 2.4 (category boundaries); (c) not applicable |
| nut-16 | Specify if nutrient intakes are reported with or without inclusion of dietary supplement intake, if applicable. | Section 2.4 (food only) |
| 17 | Other analyses: report other analyses done, e.g., analyses of subgroups and interactions and sensitivity analyses. | Sections 3.2 and 3.5; Tables 3 and S3; Figure S2 |
| nut-17 | Report any sensitivity analysis (e.g., exclusion of misreporters or outliers) and data imputation, if applicable. | Section 2.5 (extreme values reviewed, not Winsorised); Section 2.2 (ABS imputation of BMI and blood pressure) |
| **Discussion** | | |
| 18 | Key results: summarize key results with reference to study objectives. | Section 4, paragraph 1 |
| 19 | Limitations: discuss limitations of the study, taking into account sources of potential bias or imprecision. Discuss both direction and magnitude of any potential bias. | Section 4, strengths and limitations paragraph |
| nut-19 | Describe the main limitations of the data sources and assessment methods used and implications for the interpretation of the findings. | Section 4, strengths and limitations paragraph |
| 20 | Interpretation: give a cautious overall interpretation of results considering objectives, limitations, multiplicity of analyses, results from similar studies, and other relevant evidence. | Section 4, paragraphs 2–7 |
| nut-20 | Report the nutritional relevance of the findings, given the complexity of diet or nutrition as an exposure. | Section 4, paragraphs 4, 6 and 7 |
| 21 | Generalizability: discuss the generalizability (external validity) of the study results. | Section 2.1 (survey scope); Section 4, paragraph 5 |
| **Other information** | | |
| 22 | Funding: give the source of funding and the role of the funders for the present study and, if applicable, for the original study on which the present article is based. | Funding statement |
| nut-22.1 | Ethics: describe the procedure for consent and study approval from ethics committee(s). | Section 2.1 |
| nut-22.2 | Supplementary material: provide data collection tools and data as online material or explain how they can be accessed. | Data availability statement |

Items numbered 1–22 are STROBE recommendations (cross-sectional study versions shown); items marked nut- are the STROBE-nut extensions. Checklist from Lachat et al., PLoS Med 2016;13:e1002036. Section numbers refer to the main manuscript; add page and line numbers once the final layout is fixed.
BMI, body mass index; CI, confidence interval; STROBE-ME, STROBE Extension for Molecular Epidemiology.

**Table S3.** Dietary fibre intake from a single Day 1 recall and as usual intake, National Nutrition and Physical Activity Survey (NNPAS) 2011–12 and 2023.

| | Day 1 intake, estimate (95% CI) | | Usual intake, estimate (95% CI) | |
|---|---|---|---|---|
| **Group and statistic** | **2011–12** | **2023** | **2011–12** | **2023** |
| **All adults** | | | | |
| Mean (g/day) | 22.9 (22.5 to 23.3) | 20.7 (20.4 to 21.0) | 22.9 (22.5 to 23.3) | 20.8 (20.5 to 21.1) |
| 5th percentile (g/day) | 7.8 (7.4 to 8.3) | 6.7 (5.7 to 7.7) | 11.9 (11.3 to 12.5) | 10.4 (9.9 to 10.9) |
| 25th percentile (g/day) | 14.3 (14.1 to 14.6) | 13.0 (12.1 to 13.9) | 17.3 (16.8 to 17.8) | 15.6 (15.1 to 16.0) |
| Median (g/day) | 20.7 (20.2 to 21.2) | 18.7 (18.4 to 19.0) | 21.9 (21.5 to 22.3) | 19.9 (19.6 to 20.2) |
| 75th percentile (g/day) | 28.7 (28.2 to 29.2) | 26.2 (25.3 to 27.1) | 27.4 (26.9 to 27.9) | 25.1 (24.7 to 25.5) |
| 95th percentile (g/day) | 46.3 (43.9 to 48.7) | 41.4 (40.2 to 42.6) | 37.2 (36.1 to 38.2) | 34.4 (33.5 to 35.3) |
| At or above the AI (%) | 28.2 (26.8 to 29.6) | 21.7 (20.5 to 22.8) | 24.6 (22.9 to 26.4) | 17.2 (15.7 to 18.7) |
| At or above the SDT (%) | 17.0 (16.0 to 18.0) | 13.1 (12.0 to 14.1) | 11.4 (9.9 to 12.9) | 7.5 (6.4 to 8.6) |
| **Men** | | | | |
| Mean (g/day) | 24.8 (24.2 to 25.4) | 22.3 (21.9 to 22.7) | 24.5 (23.9 to 25.1) | 22.2 (21.8 to 22.6) |
| 5th percentile (g/day) | 8.7 (8.1 to 9.3) | 7.0 (6.4 to 7.6) | 13.1 (12.3 to 13.8) | 11.2 (10.5 to 11.9) |
| 25th percentile (g/day) | 15.7 (15.1 to 16.2) | 14.0 (13.1 to 14.9) | 18.7 (18.0 to 19.4) | 16.7 (16.1 to 17.2) |
| Median (g/day) | 22.4 (22.0 to 22.7) | 20.1 (19.1 to 21.1) | 23.5 (22.9 to 24.1) | 21.2 (20.8 to 21.7) |
| 75th percentile (g/day) | 31.5 (30.2 to 32.7) | 28.2 (27.3 to 29.1) | 29.2 (28.6 to 29.8) | 26.7 (26.1 to 27.3) |
| 95th percentile (g/day) | 48.2 (47.3 to 49.2) | 44.2 (42.0 to 46.4) | 39.1 (37.9 to 40.3) | 36.4 (35.2 to 37.7) |
| At or above the AI (%) | 28.6 (26.6 to 30.6) | 21.0 (19.5 to 22.6) | 22.3 (20.2 to 24.3) | 15.1 (13.4 to 16.8) |
| At or above the SDT (%) | 14.0 (12.7 to 15.3) | 9.8 (8.6 to 11.0) | 6.1 (4.7 to 7.4) | 3.7 (2.8 to 4.7) |
| **Women** | | | | |
| Mean (g/day) | 21.1 (20.6 to 21.6) | 19.2 (18.8 to 19.6) | 21.3 (20.8 to 21.9) | 19.5 (19.2 to 19.9) |
| 5th percentile (g/day) | 7.0 (6.4 to 7.7) | 6.3 (5.9 to 6.7) | 11.2 (10.5 to 11.8) | 9.9 (9.4 to 10.4) |
| 25th percentile (g/day) | 13.3 (12.9 to 13.7) | 12.3 (11.6 to 13.0) | 16.2 (15.7 to 16.7) | 14.7 (14.3 to 15.1) |
| Median (g/day) | 19.1 (18.4 to 19.8) | 17.4 (16.4 to 18.4) | 20.5 (19.9 to 21.0) | 18.7 (18.3 to 19.1) |
| 75th percentile (g/day) | 26.1 (25.1 to 27.0) | 23.9 (22.8 to 25.0) | 25.5 (24.9 to 26.1) | 23.5 (22.9 to 24.0) |
| 95th percentile (g/day) | 43.2 (38.8 to 47.6) | 37.5 (35.5 to 39.5) | 34.6 (33.4 to 35.7) | 31.9 (30.8 to 33.0) |
| At or above the AI (%) | 27.9 (26.0 to 29.8) | 22.3 (20.6 to 24.0) | 27.0 (24.4 to 29.5) | 19.3 (17.3 to 21.3) |
| At or above the SDT (%) | 19.9 (18.3 to 21.5) | 16.1 (14.6 to 17.6) | 16.5 (14.3 to 18.7) | 11.1 (9.4 to 12.7) |
| **All children** | | | | |
| Mean (g/day) | 19.7 (19.2 to 20.2) | 19.9 (19.4 to 20.5) | 19.7 (19.2 to 20.2) | 20.0 (19.5 to 20.5) |
| 5th percentile (g/day) | 7.5 (7.2 to 7.8) | 7.2 (6.2 to 8.2) | 11.6 (10.7 to 12.5) | 11.4 (10.6 to 12.3) |
| 25th percentile (g/day) | 13.2 (13.0 to 13.4) | 13.1 (12.5 to 13.7) | 15.7 (15.0 to 16.4) | 15.7 (15.1 to 16.3) |
| Median (g/day) | 18.2 (17.9 to 18.5) | 18.4 (17.6 to 19.2) | 19.1 (18.5 to 19.8) | 19.3 (18.8 to 19.8) |
| 75th percentile (g/day) | 25.0 (23.8 to 26.1) | 24.5 (23.4 to 25.6) | 23.0 (22.2 to 23.8) | 23.5 (22.7 to 24.3) |
| 95th percentile (g/day) | 37.1 (35.2 to 38.9) | 38.2 (36.1 to 40.3) | 29.6 (27.7 to 31.4) | 30.9 (29.3 to 32.6) |
| At or above the AI (%) | 42.3 (39.9 to 44.7) | 40.1 (37.3 to 42.8) | 40.9 (37.1 to 44.6) | 41.8 (38.5 to 45.0) |

Values are estimates with 95% confidence intervals in brackets. Day 1 values are observed intakes from the first recall; usual values are National Cancer Institute method estimates. Both use the Day 1 person weight, and confidence intervals were obtained with the jackknife using 60 replicate weights. Adults are aged 19 years and over and children 2–18 years.
AI, Adequate Intake; SDT, Suggested Dietary Target.

**Table S4.** Percentiles of usual dietary fibre intake (g/day), National Nutrition and Physical Activity Survey (NNPAS) 2011–12 and 2023.

| Group and percentile | 2011–12, g/day (95% CI) | 2023, g/day (95% CI) | Change, g/day (95% CI) |
|---|---|---|---|
| **All adults** | | | |
| 5th percentile | 11.9 (11.3 to 12.5) | 10.4 (9.9 to 10.9) | −1.5 (−2.3 to −0.7) |
| 25th percentile | 17.3 (16.8 to 17.8) | 15.6 (15.1 to 16.0) | −1.8 (−2.4 to −1.1) |
| 50th percentile (median) | 21.9 (21.5 to 22.3) | 19.9 (19.6 to 20.2) | −2.0 (−2.5 to −1.5) |
| 75th percentile | 27.4 (26.9 to 27.9) | 25.1 (24.7 to 25.5) | −2.3 (−3.0 to −1.7) |
| 95th percentile | 37.2 (36.1 to 38.2) | 34.4 (33.5 to 35.3) | −2.8 (−4.2 to −1.4) |
| **Men** | | | |
| 5th percentile | 13.1 (12.3 to 13.8) | 11.2 (10.5 to 11.9) | −1.8 (−2.9 to −0.8) |
| 25th percentile | 18.7 (18.0 to 19.4) | 16.7 (16.1 to 17.2) | −2.1 (−2.9 to −1.2) |
| 50th percentile (median) | 23.5 (22.9 to 24.1) | 21.2 (20.8 to 21.7) | −2.3 (−3.0 to −1.5) |
| 75th percentile | 29.2 (28.6 to 29.8) | 26.7 (26.1 to 27.3) | −2.5 (−3.4 to −1.6) |
| 95th percentile | 39.1 (37.9 to 40.3) | 36.4 (35.2 to 37.7) | −2.7 (−4.4 to −0.9) |
| **Women** | | | |
| 5th percentile | 11.2 (10.5 to 11.8) | 9.9 (9.4 to 10.4) | −1.3 (−2.1 to −0.5) |
| 25th percentile | 16.2 (15.7 to 16.7) | 14.7 (14.3 to 15.1) | −1.5 (−2.2 to −0.8) |
| 50th percentile (median) | 20.5 (19.9 to 21.0) | 18.7 (18.3 to 19.1) | −1.7 (−2.4 to −1.1) |
| 75th percentile | 25.5 (24.9 to 26.1) | 23.5 (22.9 to 24.0) | −2.0 (−2.8 to −1.2) |
| 95th percentile | 34.6 (33.4 to 35.7) | 31.9 (30.8 to 33.0) | −2.7 (−4.3 to −1.0) |
| **All children** | | | |
| 5th percentile | 11.6 (10.7 to 12.5) | 11.4 (10.6 to 12.3) | −0.1 (−1.4 to 1.1) |
| 25th percentile | 15.7 (15.0 to 16.4) | 15.7 (15.1 to 16.3) | 0.0 (−0.9 to 0.9) |
| 50th percentile (median) | 19.1 (18.5 to 19.8) | 19.3 (18.8 to 19.8) | 0.2 (−0.7 to 1.0) |
| 75th percentile | 23.0 (22.2 to 23.8) | 23.5 (22.7 to 24.3) | 0.5 (−0.6 to 1.6) |
| 95th percentile | 29.6 (27.7 to 31.4) | 30.9 (29.3 to 32.6) | 1.4 (−1.1 to 3.8) |
| **Boys** | | | |
| 5th percentile | 12.5 (11.3 to 13.6) | 12.3 (11.3 to 13.3) | −0.2 (−1.7 to 1.4) |
| 25th percentile | 16.9 (16.1 to 17.6) | 16.9 (16.1 to 17.6) | 0.0 (−1.1 to 1.1) |
| 50th percentile (median) | 20.4 (19.7 to 21.1) | 20.7 (19.9 to 21.5) | 0.3 (−0.8 to 1.4) |
| 75th percentile | 24.4 (23.4 to 25.3) | 25.1 (23.9 to 26.2) | 0.7 (−0.8 to 2.2) |
| 95th percentile | 31.1 (28.9 to 33.3) | 32.8 (30.8 to 34.8) | 1.7 (−1.3 to 4.7) |
| **Girls** | | | |
| 5th percentile | 11.0 (10.0 to 11.9) | 10.8 (9.8 to 11.8) | −0.2 (−1.6 to 1.2) |
| 25th percentile | 14.8 (14.0 to 15.6) | 14.7 (13.9 to 15.4) | −0.1 (−1.2 to 1.0) |
| 50th percentile (median) | 17.9 (17.2 to 18.6) | 17.9 (17.2 to 18.6) | 0.0 (−1.0 to 1.0) |
| 75th percentile | 21.4 (20.6 to 22.3) | 21.7 (20.8 to 22.5) | 0.2 (−0.9 to 1.4) |
| 95th percentile | 27.4 (25.2 to 29.6) | 28.0 (26.6 to 29.5) | 0.6 (−2.0 to 3.2) |

Values are estimates with 95% confidence intervals in brackets, estimated with the National Cancer Institute method; confidence intervals were obtained with the jackknife using 60 replicate weights. Changes are 2023 minus 2011–12; negative values indicate a decrease.

#### Supplementary Figures

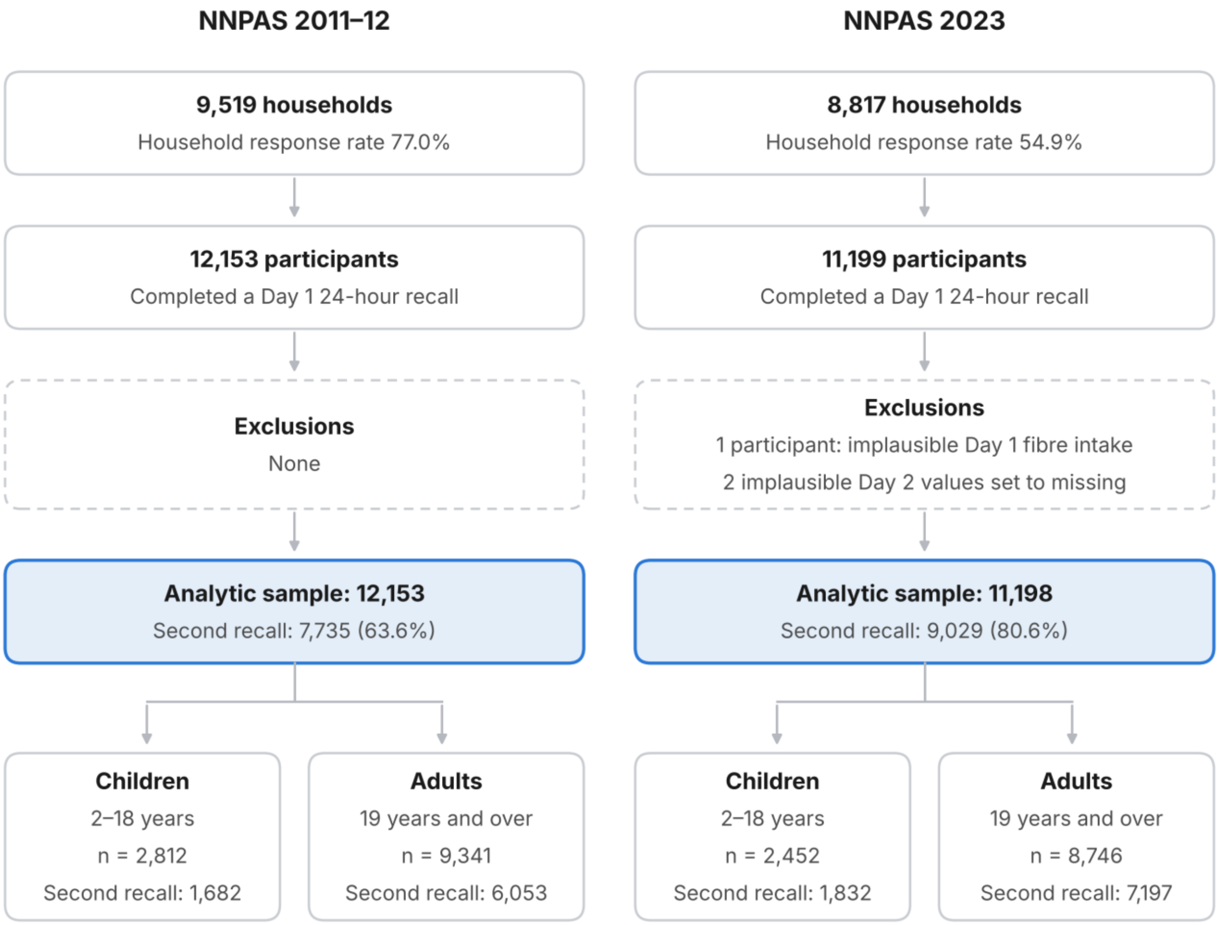


**Figure S1.** Flow of participants in the National Nutrition and Physical Activity Survey (NNPAS) 2011–12 and 2023. Percentages for the second recall are of the analytic sample. Implausible values were identified by review of extreme intakes (Section 2.5).

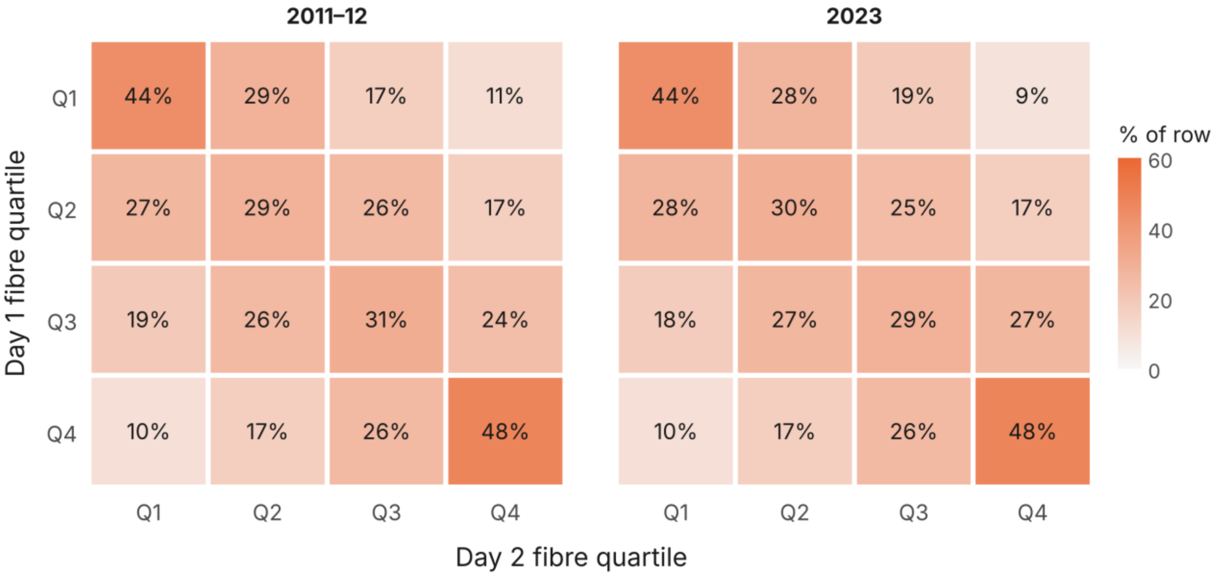


**Figure S2.** Agreement between Day 1 and Day 2 fibre intake quartiles in adults aged 19 years and over with two recalls, NNPAS 2011–12 and 2023. Adults with two recalls (2011–12, n = 6,053; 2023, n = 7,197), weighted with the Day 1 person weight. Each row shows the percentage of adults in a Day 1 quartile who fell in each Day 2 quartile; rows sum to 100%. Correlation between Day 1 and Day 2 intake: r = 0.43 (2011–12) and r = 0.45 (2023).
